# A metabolite index of ultra-processed food intake is associated with stroke, cancer mortality, and all-cause mortality

**DOI:** 10.64898/2026.01.09.26343724

**Authors:** Zsuzsanna Ament, Jonathan Duskin, Amelia Demopoulos BA, Varun M. Bhave, Ana-Lucia Garcia Guarniz, Carol R. Oladele, Suzanne Judd, Hemant K. Tiwari, Laura C. Pinheiro, M. Ryan Irvin, Douglas E. Levy, Anne N. Thorndike, W. Taylor Kimberly

## Abstract

Ultra-processed foods (UPFs) are associated with adverse health outcomes, but measurement of UPF intake in epidemiological studies remains challenging. Dietary assessments typically employ recall questionnaires, which may lead to measurement errors and misclassifications. Here, we develop a plasma metabolite index of UPF consumption to objectively measure UPF intake. Using two years of authentic food purchasing records from the ChooseWell 365 cohort, we identify metabolites associated with long-term UPF intake. We then apply the UPF-metabolite index to the REasons for Geographic and Racial Differences in Stroke (REGARDS) cohort, a population-based cohort of U.S. adults followed for incident health outcomes. The UPF-metabolite index was associated with leading causes of morbidity and mortality, including incident stroke, all-cause mortality, and cancer mortality. Mediation analysis revealed that the UPF-metabolite index accounts for 62% of the association between UPF intake and stroke risk. These findings suggest that the UPF-metabolite index offers an objective method for assessing UPF intake and its contribution to diet-related disease risk.

**Teaser:** Plasma-based markers of ultra-processed foods (UPFs) predict disease risk, including stroke, cancer mortality, and all-cause mortality.

## Introduction

Ultra-processed foods (UPFs) have become a dominant component of modern diets and are associated with an increased risk of cardiometabolic disease, stroke, and cognitive decline (*1–5*). Frequently classified based on the NOVA system, foods are categorized into four food groups based on the extent and type of processing, ranging from unprocessed (NOVA 1) to ultra-processed (NOVA 4) (*6*, *7*). Items in the NOVA 4 category typically include ingredients derived from industrial processes and often contain multiple additives designed to augment palatability and extend shelf life (*8*, *9*).

Despite growing evidence supporting the importance of UPFs in nutritional and health policy, (*10*, *11*) accurately assessing of UPF intake remains challenging. Classifying foods into NOVA categories is particularly difficult in epidemiological studies, which serve as the primary sources of insight into the health effects of UPF consumption (*1*, *2*, *12–14*). These studies are essential for unraveling associations between UPF intake and long-term health outcomes through large-scale, longitudinal observation. However, food intake is often assessed through cross-sectional, self-reported food frequency questionnaires, which lack adequate detail on specific food types and their constituent ingredients for precise UPF classification. This limitation introduces variability across studies, which can affect the accuracy and comparability of associations between UPF consumption and health outcomes.

To address the challenges of accurately measuring UPF intake in epidemiological studies, we hypothesized that UPF consumption could be objectively captured through blood-based biomarkers. Given that food intake influences host metabolism, metabolomic biomarkers are well-suited to capture and reflect UPF consumption. This hypothesis is further supported by prior studies that have shown that 1) circulating metabolites are associated with several other dietary patterns (*15–17*), and 2) specific metabolites have been associated with a range of disease outcomes, including stroke (*18–20*), cardiometabolic disease (*21*, *22*), and cancer (*23*, *24*).

In this study, we tested the hypothesis that circulating metabolites would serve as objective biomarkers of UPF consumption and subsequent health outcomes. We first developed a UPF-metabolite index using detailed food purchase data, dietary recall surveys, and plasma samples from the ChooseWell 365 cohort (*25–27*). We then leveraged the REasons for Geographic and Racial Differences in Stroke (REGARDS) study (*14*, *28–31*), a large, geographically dispersed U.S. cohort of community-dwelling adults with baseline plasma samples, who have been followed for more than 15 years for a variety of health outcomes. We evaluated associations between the UPF-metabolite index and the primary outcome of interest in REGARDS, incident stroke. We then examined whether the UPF-metabolite index mediated the relationship between high UPF intake and incident stroke. Finally, we evaluated additional outcomes, including heart failure, myocardial infarction events, as well as all-cause, cancer, and cardiovascular mortality. Given that stroke, cardiovascular disease, and cancer are among the leading causes of death and disability worldwide, understanding dietary contributions to these outcomes remains a critical public health priority.

## Results

### Development of the UPF-metabolite index in the ChooseWell 365 study

We first characterized UPF consumption in the ChooseWell 365 study, which included 470 individuals with plasma samples drawn at the two-year follow-up visit (Table 1). This cohort was predominantly White (81.7%), female (79.6%), and had a mean age of 44 ± 12 years. UPF intake was quantified from detailed food purchasing records collected over a two-year period. A total of 2,662 food items with distinct identifiers were categorized based on the NOVA classification system (*32*). Cafeteria purchases (*N*=931,614 separate purchase records) were aggregated by participant, and UPF intake was determined as the percentage of total calories purchased. Participants purchased an average of 46.8% ± 17.4% of total calories from UPFs. Overall, UPF purchases varied by demographic and clinical characteristics, consistent with prior reports (*33*) (Supplemental Figure S1).

**Table 1.** Characteristics of the ChooseWell 365 participants.

| **Characteristic** | **Overall (N=470)** |
| --- | --- |
| **Demographics** |  |
| Age, years (mean ± SD) | 44 ± 12 |
| Female, N (%) | 376 (80%) |
| **Race, N (%)** |  |
| White | 384 (81.7%) |
| Black | 42 (8.9%) |
| Other* | 44 (9.4%) |
| **Diet proportion (% calories ± SD)** |  |
| NOVA 1 | 39.6 ± 17.6% |
| NOVA 2 | 0 ± 0% |
| NOVA 3 | 13.5 ± 8.4% |
| NOVA 4 | 46.7 ± 17.4% |

Values are presented as *N* (%) for categorical variables and mean ± standard deviation (SD) for continuous variables. NOVA classification categories represent food processing levels: NOVA 1 (unprocessed/ minimally processed), NOVA 2 (processed culinary ingredients), NOVA 3 (processed foods), and NOVA 4 (ultra-processed foods).
\*Other race categories included Asian (*N*=22, 4.7%), those with more than one race (*N*=6, 1.3%), Native Hawaiian/ Pacific Islander (*N*=1, 0.2%), and participants who preferred not to answer (*N*=15, 3.2%).

We next identified plasma metabolites associated with UPF purchases. Out of 160 plasma metabolites measured, a total of 14 were associated with average UPF intake during the two-year study. Sensitivity analyses, which accounted for possible alternative classification of ambiguous food items, confirmed 10 robust associations. Among the 10 confirmed metabolites, four were positively associated with higher UPF consumption, and six metabolites were inversely associated (Table 2).

**Table 2:**
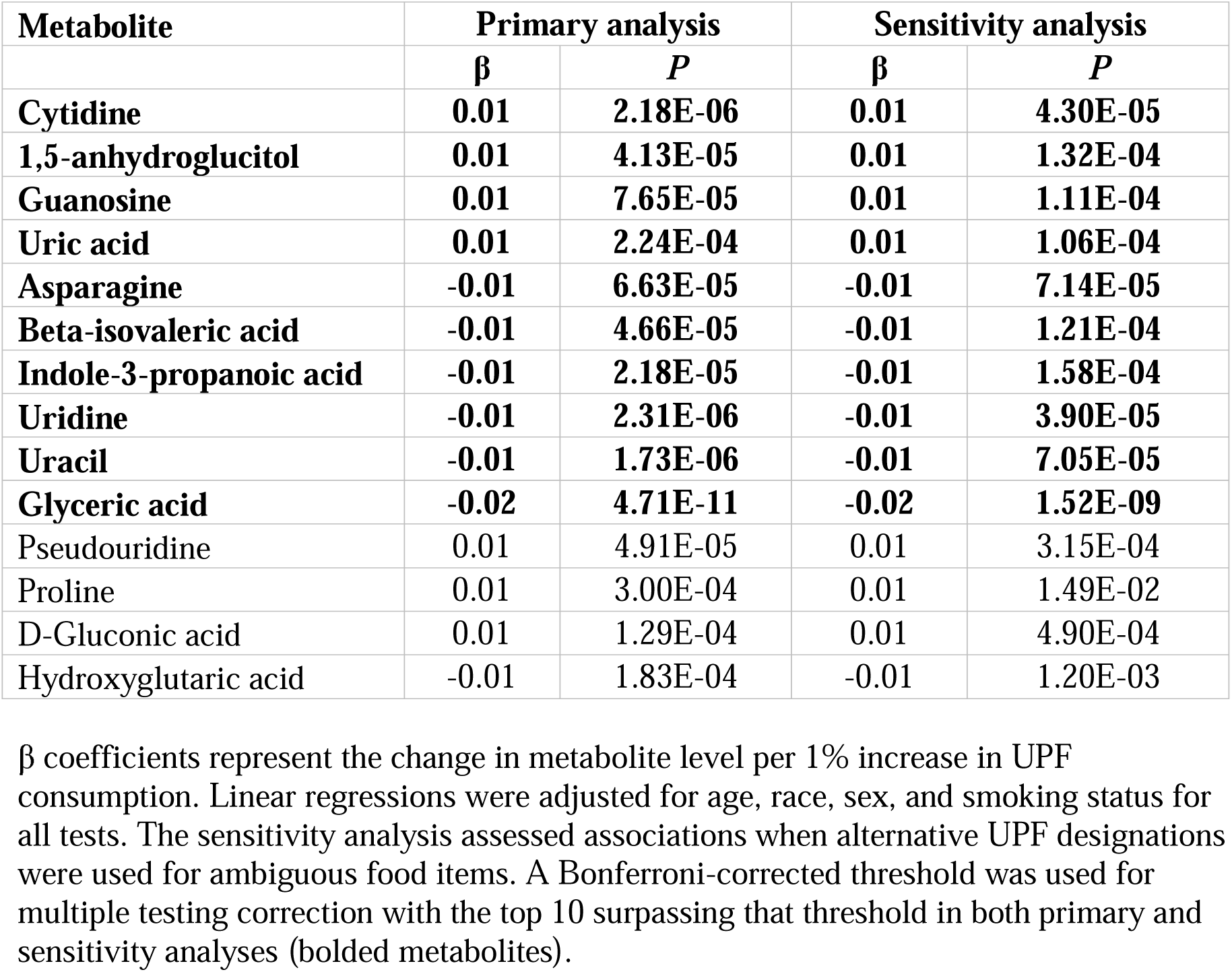
Metabolite associations with UPF intake in the ChooseWell 365 study.

| Metabolite | Primary analysis |  | Sensitivity analysis |  |
| --- | --- | --- | --- | --- |
| | $\beta$ | *P* | $\beta$ | *P* |
| **Cytidine** | **0.01** | **2.18E-06** | **0.01** | **4.30E-05** |
| **1,5-anhydroglucitol** | **0.01** | **4.13E-05** | **0.01** | **1.32E-04** |
| **Guanosine** | **0.01** | **7.65E-05** | **0.01** | **1.11E-04** |
| **Uric acid** | **0.01** | **2.24E-04** | **0.01** | **1.06E-04** |
| **Asparagine** | **-0.01** | **6.63E-05** | **-0.01** | **7.14E-05** |
| **Beta-isovaleric acid** | **-0.01** | **4.66E-05** | **-0.01** | **1.21E-04** |
| **Indole-3-propanoic acid** | **-0.01** | **2.18E-05** | **-0.01** | **1.58E-04** |
| **Uridine** | **-0.01** | **2.31E-06** | **-0.01** | **3.90E-05** |
| **Uracil** | **-0.01** | **1.73E-06** | **-0.01** | **7.05E-05** |
| **Glyceric acid** | **-0.02** | **4.71E-11** | **-0.02** | **1.52E-09** |
| Pseudouridine | 0.01 | 4.91E-05 | 0.01 | 3.15E-04 |
| Proline | 0.01 | 3.00E-04 | 0.01 | 1.49E-02 |
| D-Gluconic acid | 0.01 | 1.29E-04 | 0.01 | 4.90E-04 |
| Hydroxyglutaric acid | -0.01 | 1.83E-04 | -0.01 | 1.20E-03 |

$\beta$ coefficients represent the change in metabolite level per 1% increase in UPF consumption. Linear regressions were adjusted for age, race, sex, and smoking status for all tests. The sensitivity analysis assessed associations when alternative UPF designations were used for ambiguous food items. A Bonferroni-corrected threshold was used for multiple testing correction with the top 10 surpassing that threshold in both primary and sensitivity analyses (bolded metabolites).

A composite UPF-metabolite index was derived from the 10 metabolites, identified using models with minimal adjustment (age, race, sex, and smoking) to avoid over-adjustment bias, as clinical comorbidities may lie on the causal pathway between UPF intake and outcome. This index, as expected, was associated with overall UPF intake (β = 6.67; 95% CI: 5.50–7.84; *P* = 6.14 × 10 ^26^). We additionally assessed the index by testing its association with self-reported UPF intake from a 24-hour dietary recall conducted at the end of the study. Each standard deviation increase in the UPF-metabolite index corresponded to 3.01% higher overall UPF consumption (95% CI: 2.05–3.97; *P* = 1.44 × 10 ^9^). As a sensitivity analysis, we tested the association after adjustment for comorbidities including hypertension, prior stroke or TIA, diabetes, coronary heart disease, cancer, and smoking (β = 2.90; 95% CI: 1.88–3.92; *P* = 3.29 × 10). The association stayed robust, demonstrating that the metabolite index captures UPF-related biological signatures independent of prevalent health conditions. Additionally, we evaluated associations between the UPF-metabolite index and prevalent cardiometabolic conditions within the ChooseWell 365 study population, and found that the UPF-metabolite index was associated with diabetes, obesity, hypertension, and hyperlipidemia (Figure 1a–d).

**Fig. 1.**
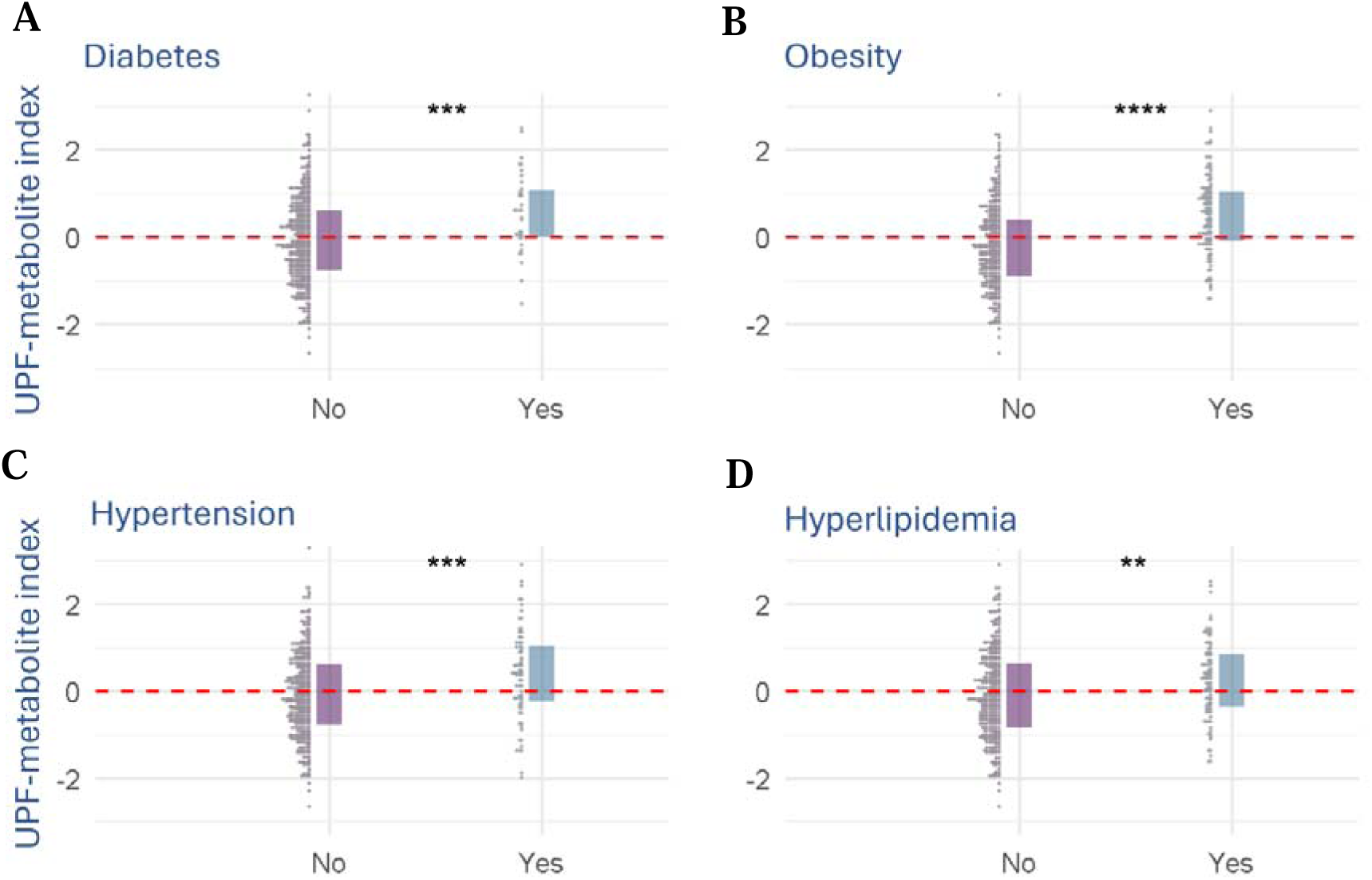
Distributional differences in the UPF-metabolite index by health status in the ChooseWell 365 study. Distribution of the UPF-metabolite index stratified by presence or absence of **(A)** diabetes, **(B)** obesity, **(C)** hypertension, and **(D)** hyperlipidemia in ChooseWell 365 study participants (*N*=470). Plots illustrate median, interquartile range, and individual data points. (**P<0.01, ***P<0.001, ****P<0.0001).

### Validation of the UPF-metabolite index in the REGARDS study

We next validated the UPF-metabolite index in the REGARDS study, using an existing stroke case-cohort design that included 2,165 participants (*18*) (Table 3). In the random cohort sample, the mean age was 64 ± 9 years, 55% were female, and 32% were Black participants. Among stroke cases, the mean age was 70 ± 9 years, 48% were female, and 35% were Black participants. Based on food frequency questionnaire data, REGARDS participants consumed an average of 48% ± 12% of daily calories from UPFs (*15*), similar to the percentage of UPF intake among Choosewell 365 participants (Supplementary Figure 2).

**Table 3:** Baseline characteristics of the REGARDS stroke case-cohort study.

|  | **Random**<br>**subgroup**<br>**(N=1056)** | **Stroke cases**<br>**(N=1109)** | ***P*** |
| --- | --- | --- | --- |
| **Demographics** |  |  |  |
| Age, years (mean ± SD) | 64.3 ± 9.0 | 69.8 ± 8.7 | <0.001 |
| Female (%) | 55.3 | 47.5 | <0.001 |
| White | 65.4 | 68 | 0.124 |
| **Clinical Measures (%)** |  |  |  |
| Current smoker | 13.8 | 14.9 | 0.405 |
| Hypertension | 70.2 | 82.7 | <0.001 |
| Diabetes mellitus | 17.2 | 24.6 | <0.001 |
| Cardiovascular disease | 15.0 | 27.2 | <0.001 |
| Left ventricular hypertrophy | 7.4 | 17.5 | <0.001 |
| **Atrial fibrillation** | 8.0 | 13.8 | <0.001 |
| **Daily diet proportion**<br>**(% calories ± SD)** |  |  |  |
| NOVA 1 | 38.7 ± 11.6% | 38.2 ± 11.6% | 0.311 |
| NOVA 2 | 5.2 ± 4.7 | 4.8 ± 4.6 | 0.027 |
| NOVA 3 | 8.3 ± 6.1 | 8.5 ± 6.8 | 0.419 |
| NOVA 4 | 47.9 ± 11.9 | 48.5 ± 12.3 | 0.180 |
| **Daily diet proportion**<br>**(% grams ± SD)** |  |  |  |
| NOVA 1 | 73.9 ± 13.2% | 72.8 ± 13.5% | 0.046 |
| NOVA 2 | 0.6 ± 0.7 | 0.6 ± 0.7 | 0.269 |
| NOVA 3 | 5.9 ± 6.6 | 6.2 ± 7.5 | 0.355 |
| NOVA 4 | 19.6 ± 12.2 | 20.4 ± 12.4 | 0.094 |

Data presented as percentage or mean ± standard deviation (SD). Percentages are survey-weighted to account for the case-cohort sampling design, representing the distribution in the full REGARDS cohort ( $N=30,239$ ). *P*-values are from survey-weighted chi-square tests for categorical variables and t-tests for continuous variables, accounting for the case-cohort design. NOVA classification categories represent food processing levels: NOVA 1 (unprocessed/minimally processed), NOVA 2 (processed culinary ingredients), NOVA 3 (processed foods), and NOVA 4 (UPFs).

In the REGARDS cohort, the UPF-metabolite index was associated with overall dietary UPF consumption, whether expressed as a percentage of calories (β = 3.1; 95% CI: 2.5–3.7; *P* = 2.9×10 ^23^) or as a percentage of grams consumed (β = 2.3; 95% CI: 1.7–3.0; *P* = 2.2×10 ^12^; Supplementary Figure 3). The UPF-metabolite index was also associated with prevalent comorbidities, including obesity, diabetes, hypertension, and hyperlipidemia (Supplementary Figure 4). Individual metabolite associations with overall UPF intake for both cohorts are provided as Supplementary Table 1.

## UPF-metabolite index and incident health outcomes

### Associations between the UPF-metabolite index and incident stroke

We next examined associations between the UPF-metabolite index and future health outcomes, first focusing on incident stroke since this is the primary outcome of interest in the REGARDS study. In a minimally adjusted Cox proportional hazards model, each standard deviation increase in the UPF-metabolite index was associated with a nearly two-fold higher risk of stroke (HR = 1.93; 95% CI: 1.64–2.27; *P* = 9.01×10 ¹) and this association remained after adjustment for stroke-related risk factors (HR = 1.53; 95% CI: 1.29–1.81; *P* = 1.32×10 ^6^; Table 4).

**Table 4:** Incident stroke associations with UPF-metabolite index in the REGARDS study.

| **Model** | **HR** | **95% CI** | ***P*** |
| --- | --- | --- | --- |
| UPF-metabolite index (Base model) | 1.27 | 1.20 □ 1.35 | 9.01E-16 |
| UPF-metabolite index (Fully adjusted model) | 1.17 | 1.10 □ 1.24 | 1.32E-06 |

Base-adjusted Cox proportional hazards models included age, race, sex, and age by race interaction as covariates. Fully adjusted models additionally included stroke-related vascular risk factors: smoking status, left ventricular hypertrophy, atrial fibrillation, hypertension, diabetes, and coronary artery disease.

To evaluate a potential dose–response relationship, we divided the UPF-metabolite index into quartiles and further examined these in fully adjusted models. Median UPF intake increased across quartiles, from 43.3% in Q1 to 53.2% in Q4. Relative to the lowest quartile, the risk for stroke was elevated in the highest quartile (Q4: HR = 1.49; 95% CI: 1.26–1.77; *P* = 4.45 x 10^-6^) but not the second or third quartile, suggesting a threshold effect with risk primarily among individuals in the top quartile of the UPF-metabolite index (Figure 2).

**Fig 2.**
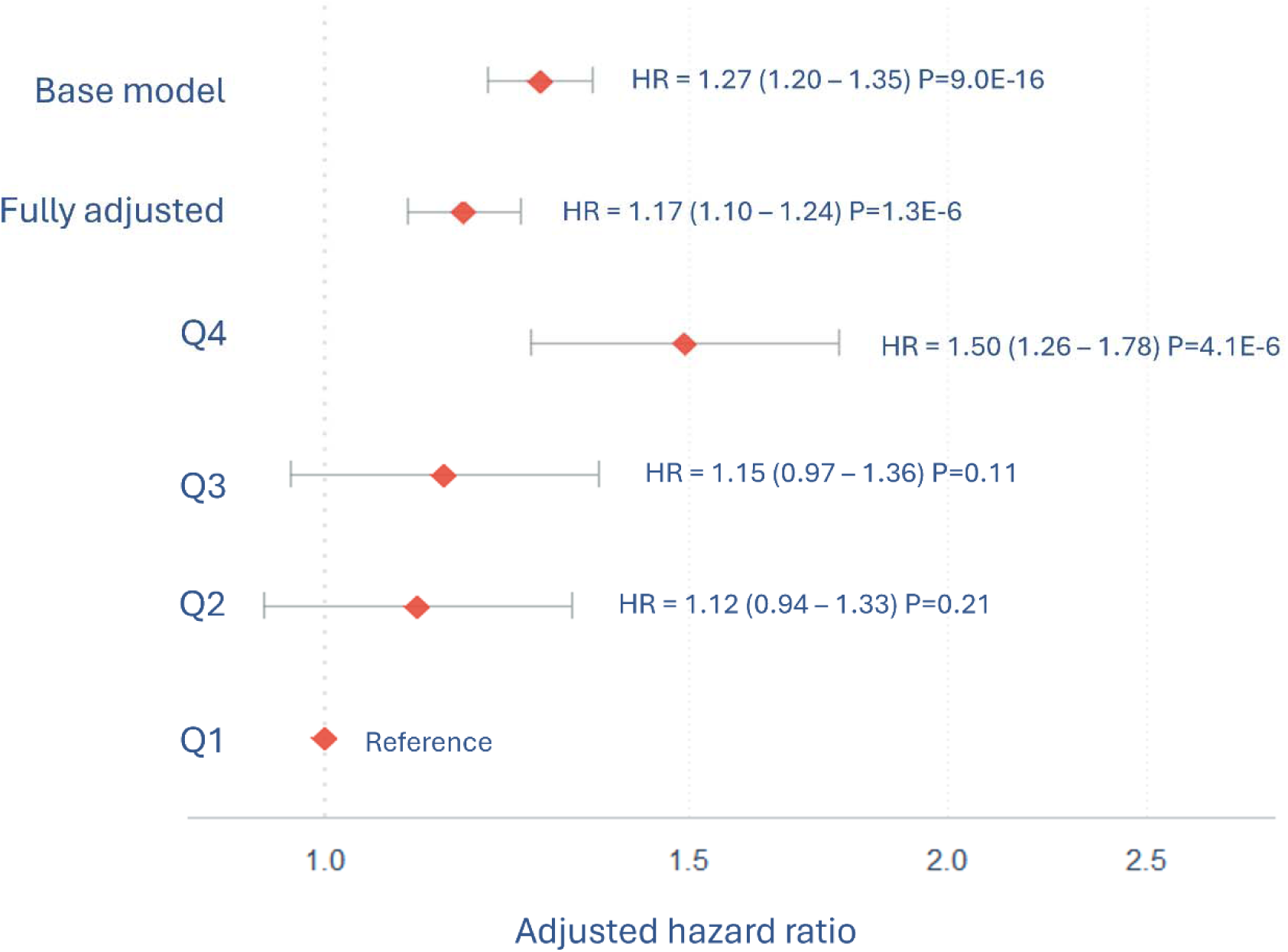
The UPF-metabolite index is associated with incident stroke. The forest plot displays hazard ratios (HR) and 95% confidence intervals for stroke risk per one standard deviation (SD) increase in the UPF-metabolite index (continuous models) and by quartiles (Q1-Q4, with Q1 as reference). Base models were adjusted for age, race, gender, and age by race interaction. Fully adjusted models additionally included smoking status, left ventricular hypertrophy, atrial fibrillation, hypertension, diabetes mellitus, and coronary artery disease. The model using quartiles was adjusted for covariates similar to the full model. Analysis used Cox proportional hazards models with the Breslow method for tied event times, weighted for the case-cohort sampling design.

To further characterize the role of the UPF-metabolite index on incident stroke, we conducted mediation analysis to determine whether the UPF-metabolite index mediated the relationship between UPF dietary intake and incident stroke risk (Figure 3). To satisfy the conditions for mediation analysis, we confirmed that overall UPF intake as a percentage of total caloric intake was associated with an elevated UPF-metabolite index (OR = 3.63; 95% CI: 2.60–4.79; *P* = 4.19×10 ^16^), and that an overall higher UPF intake was associated with an increased risk of incident stroke (HR= 1.17; 95% CI: 1.02–1.34; *P* = 0.024). The mediating effect of the UPF-metabolite index on the relationship between UPF dietary intake and stroke risk was β = 0.10 (*P* = 0.050), accounting for 62% of the total effect.

**Fig 3.**
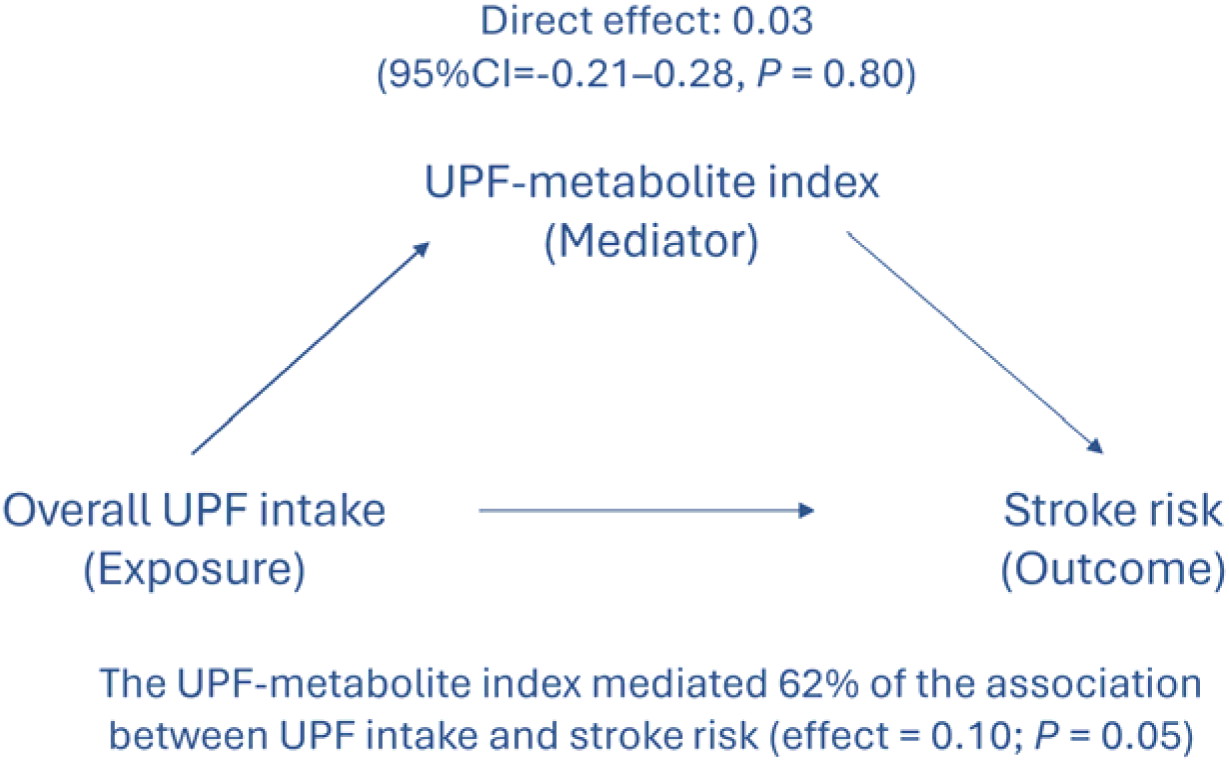
Mediation of the association between UPF intake and stroke risk by the UPF-metabolite index. Inverse odds ratio weighting (IORW) mediation analysis showing that the UPF-metabolite index mediated the association between UPF dietary intake and incident stroke (β = 0.10; *P* = .050), accounting for 62% of the total effect. Models were adjusted for age, race, sex, and age-by-race interaction.

### Associations between the UPF-metabolite index and other health outcomes

To explore associations with other health outcomes, we evaluated additional endpoints from the REGARDS study in models adjusted for demographic characteristics, and in fully adjusted models that additionally included comorbidities (*34*, *35*). This analysis was focused on the random cohort sample, and mediation analysis was not performed due to a lack of association between overall UPF intake and the additional endpoints, in this smaller sample set of the random cohort participants. Figure 4 presents a forest plot of the associations between the UPF-metabolite index and major health outcomes, adjusted for both demographic factors and comorbidities. Table 5 includes results from both base-adjusted and fully adjusted models. In the fully adjusted models, the UPF-metabolite index was significantly associated with all-cause mortality (HR = 1.26 per SD; 95% CI: 1.12–1.42; *P* = 1.38×10 ^4^), and cancer mortality (HR = 1.33 per SD; 95% CI: 1.02–1.74; *P* = 3.80×10^-2^). Cardiovascular death and heart failure, but not myocardial infarction, was associated with the UPF-metabolite index in base-adjusted models. However, these associations were attenuated after adjustments for covariates.

**Fig 4.**
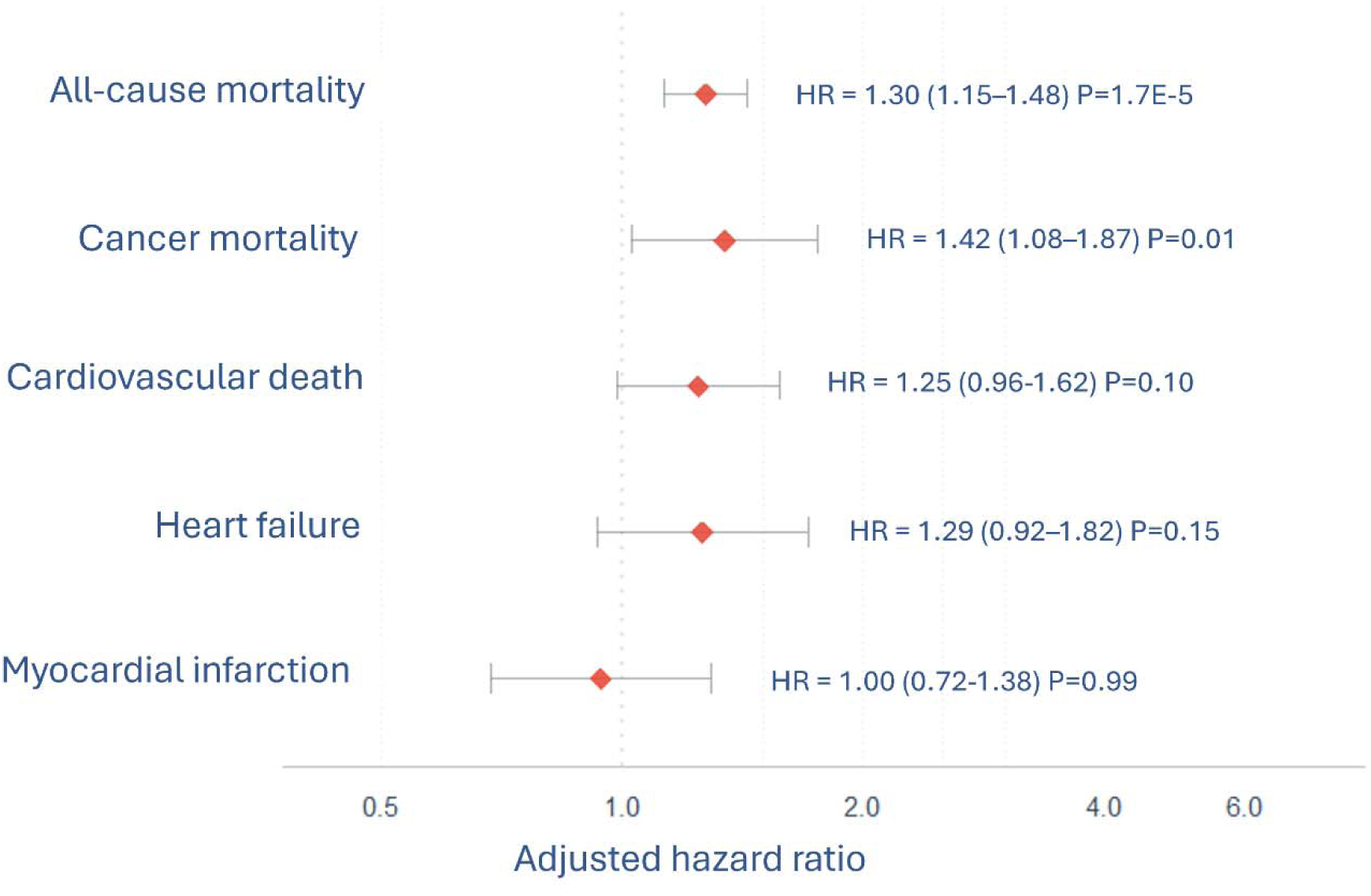
Association between UPF-metabolite index and multiple health outcomes in the REGARDS study random cohort sample. The forest plot displays adjusted hazard ratios (HR) and 95% confidence intervals for various health outcomes per one standard deviation (SD) increase in the UPF-metabolite index. Models were adjusted for age, race, sex, smoking status, body mass index, hypertension, diabetes mellitus, dyslipidemia, history of myocardial infarction, and history of cancer. Analysis used Cox proportional hazards models with the Breslow method for tied event times, restricted to participants in the random cohort (*N*=1,056).

**Table 5:**
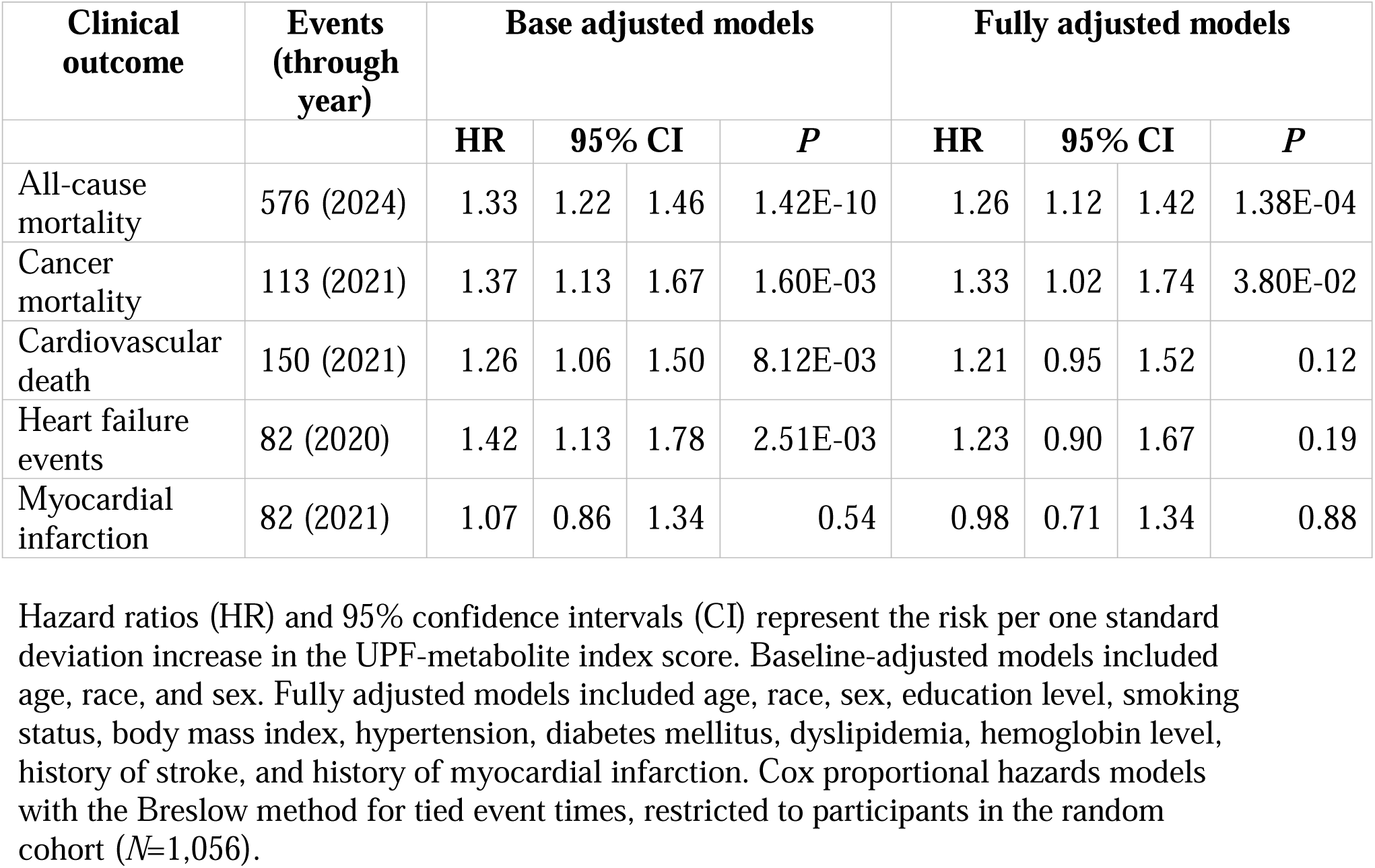
Association of UPF-metabolite index with additional clinical outcomes in the REGARDS study.

| Clinical outcome | Events (through year) | Base adjusted models |  |  |  | Fully adjusted models |  |  |  |
| --- | --- | --- | --- | --- | --- | --- | --- | --- | --- |
|  |  | HR | 95% CI |  | *P* | HR | 95% CI |  | *P* |
| All-cause mortality | 576 (2024) | 1.33 | 1.22 | 1.46 | 1.42E-10 | 1.26 | 1.12 | 1.42 | 1.38E-04 |
| Cancer mortality | 113 (2021) | 1.37 | 1.13 | 1.67 | 1.60E-03 | 1.33 | 1.02 | 1.74 | 3.80E-02 |
| Cardiovascular death | 150 (2021) | 1.26 | 1.06 | 1.50 | 8.12E-03 | 1.21 | 0.95 | 1.52 | 0.12 |
| Heart failure events | 82 (2020) | 1.42 | 1.13 | 1.78 | 2.51E-03 | 1.23 | 0.90 | 1.67 | 0.19 |
| Myocardial infarction | 82 (2021) | 1.07 | 0.86 | 1.34 | 0.54 | 0.98 | 0.71 | 1.34 | 0.88 |

Hazard ratios (HR) and 95% confidence intervals (CI) represent the risk per one standard deviation increase in the UPF-metabolite index score. Baseline-adjusted models included age, race, and sex. Fully adjusted models included age, race, sex, education level, smoking status, body mass index, hypertension, diabetes mellitus, dyslipidemia, hemoglobin level, history of stroke, and history of myocardial infarction. Cox proportional hazards models with the Breslow method for tied event times, restricted to participants in the random cohort ( $N=1,056$ ).

## Discussion

Our study identified a robust metabolite biomarker signature of UPF consumption that is associated with incident stroke, all-cause mortality, and cancer mortality. Using targeted metabolomics in two independent cohorts with complementary strengths, including objective food purchasing records in ChooseWell 365 study and adjudicated health outcomes in the REGARDS study population, we identified 10 plasma metabolites that were consistently associated with overall UPF intake. We developed a composite UPF-metabolite index from these 10 metabolites, and in the REGARDS stroke cohort, we found that the UPF-metabolite index was strongly associated with incident stroke. Mediation analysis revealed that the UPF-metabolite index accounted for approximately 62% of the total effect of UPF intake on stroke risk, suggesting that this metabolomic signature captures key biological pathways linking UPF consumption and stroke. Although stroke was the primary outcome of interest, we assessed additional associations between the UPF-metabolite index and observed associations with all-cause mortality and cancer mortality in the REGARDS random cohort. In contrast to prior studies demonstrating associations between UPF consumption and cardiovascular disease (*4*), our metabolite-based UPF index did not show strong associations with myocardial infarction, heart failure, or cardiovascular death. These differences, both within our analysis, and in comparison to prior literature (*4*), may reflect limited statistical power due to the smaller number of events in the random cohort, or greater heterogeneity in the underlying pathophysiology of these outcomes.

The UPF-metabolite index may offer advantages relative to traditional dietary assessment methods for studying UPF consumption in epidemiological research. Metabolomic biomarkers provide a more objective measure of dietary exposure compared to self-reported dietary reports, and reflect both food intake and individual metabolic responses. The UPF-metabolite index could serve as a valuable addition to complement dietary questionnaires in large-scale observational studies, potentially reducing measurement error and improving the precision of diet-disease associations. From a clinical perspective, metabolite-based biomarkers may also enhance patient motivation by providing measurable shorter-term targets that can be achieved within years rather than decades, while still contributing to the prevention of long-term health outcomes. Furthermore, the metabolomic approach may be particularly useful in clinical trials evaluating interventions to reduce UPF consumption, where objective biomarkers could serve as endpoints or intermediate measures of dietary change. Our findings are in line with a recent study, which similarly developed poly-metabolite scores predictive of UPF intake using metabolomics data and validated them in a controlled feeding trial (*36*), although the sample size was smaller and health outcomes were not included in that study.

The individual metabolites identified in this study also provide insights into potential biological pathways that link UPF consumption to disease outcomes. For example, there was an inverse association (e.g., a protective effect) with indole-3-propionic acid, a microbiota-derived metabolite which, aligned with studies (*37*), suggests that UPF consumption may disrupt beneficial gut microbial metabolism. The positive associations with cytidine and guanosine indicate alterations in nucleoside metabolism, while elevated uric acid and 1,5-anhydroglucitol may reflect oxidative stress associated with high sugar intake, a common added ingredient in UPFs. Negative associations with uridine and beta-isovaleric acid may reflect increased metabolic stress or reduced availability of protective metabolic intermediates. However, further mechanistic studies are needed to establish causal relationships between these metabolites, the potential role of the gut microbiome, and subsequent health outcomes.

An important strength of our approach is that the UPF-metabolite index reflects long-term dietary patterns rather than shorter-term fluctuations, which more commonly affect controlled feeding trials. In the ChooseWell 365 study, we derived the index from food purchasing data collected over two years, which likely captures habitual dietary behaviors that are most relevant to chronic disease risk. Similarly, in REGARDS, the index was associated with dietary patterns assessed at baseline and predicted health outcomes over a decade of follow-up. This temporal stability is crucial for epidemiological studies examining diet-disease relationships, as long-term dietary exposures are more likely to influence chronic disease development rather than brief dietary changes.

However, limitations should be noted. First, daily dietary intake outside the cafeteria in ChooseWell 365 was not systematically recorded, and dietary assessment in REGARDS relied on food frequency questionnaires, each of which may be subject to some bias. Second, while mediation analyses strengthen the plausibility of causality for these findings, experimental studies are needed to confirm causation. Third, the index reflects a combination of metabolites with potentially heterogeneous biological roles, and further work is needed to decompose individual contributions. Future studies should explore how UPF-associated metabolic changes interact with microbiome composition, genetic susceptibility, and lipid metabolism. Intervention studies assessing whether reducing UPF intake can reverse these metabolic signatures and whether reformulated processed foods can attenuate these effects could inform dietary policy and food-based innovation. Finally, there are different metabolomic methods detecting different sets of metabolites. Future studies should seek to harmonize metabolite coverage to ensure greater consistency across cohorts.

These findings are supported across diverse populations, as the ChooseWell 365 study consisted predominantly of White female participants, and the REGARDS study included both Black and White participants, who were older and had a higher burden of comorbidities. The consistency of metabolite associations across these different populations suggests robustness. However, further studies in populations with differing demographic, health, and dietary profiles would further strengthen generalizability.

In conclusion, our study identified and validated a metabolic signature of ultra-processed food consumption that was strongly associated with incident stroke, all-cause mortality, and cancer mortality. This UPF-metabolite index may serve as a sensitive biomarker of dietary quality and metabolic health, while offering insight into the biological pathways through which food processing contributes to chronic disease risk. These findings support the utility of metabolomics in advancing precision nutrition and guiding evidence-based public health policies concerning UPF consumption.

## Materials and Methods

### Experimental Design

The primary objective of this study was to identify plasma metabolite markers associated with UPF intake and to construct a composite UPF-metabolite index that could serve as an objective biomarker of UPF consumption. The secondary objectives included external validation of the index in an independent cohort, and evaluation of its associations with major health outcomes, including incident stroke, cardiovascular diseases, cancer and all-cause mortality.

The study was conducted in two phases across two distinct cohorts. In the discovery phase, we used the ChooseWell 365 cohort, which included detailed food purchasing data and plasma samples collected at a two-year follow-up visit. UPF intake was quantified using the NOVA classification system applied to cafeteria purchases, and plasma metabolites were profiled using liquid chromatography–tandem mass spectrometry (LC-MS/MS). We identified individual metabolites associated with UPF intake and developed a composite UPF-metabolite index to reflect overall dietary exposure.

In the validation phase, the UPF-metabolite index was assessed in the REGARDS study, a U.S.-based cohort of geographically dispersed, community-dwelling participants. Metabolomics data were available for participants in the stroke case-cohort sub-study, along with adjudicated health outcomes. We evaluated associations between the UPF-metabolite index and incident stroke, and examined other clinical endpoints, including cardiovascular events and cause-specific mortality.

### Study cohorts

#### ChooseWell 365 study

The ChooseWell 365 study was used as the discovery cohort. This 24-month randomized clinical trial was conducted among employees at Massachusetts General Hospital and originally designed to evaluate a behavioral intervention to promote healthy food choices and prevent weight gain (*26*). The methods and design of the ChooseWell 365 study has been described in detail elsewhere (*25*, *26*, *38*, *39*). Briefly, between September 2016 and February 2018, the study enrolled 602 subjects. To be eligible for participation, employees needed to regularly use the hospital cafeteria, as defined by cafeteria badge use, at least 4 times per week during the 12 weeks prior to enrollment. Key exclusion criteria included plans to leave employment within one-year, current pregnancy, a history of eating disorders, recent or planned weight loss surgery, existing enrollment in weight loss programs, or employment in the cafeteria or the study’s clinical research facility. Of the original 602 participants, 470 (78.1%) had a plasma sample at the two-year follow-up time point available for metabolomic analyses. Detailed individual food purchases were examined over the full two-year period from these 470 participants. In addition to plasma sample collection, dietary intake was assessed at study completion using a 24-hour recall survey. This design enabled us to examine associations between long-term UPF intake and plasma metabolite levels, and to internally validate findings by comparison with dietary recall-based methods.

#### REGARDS study

The REGARDS study served as the validation cohort and to investigate associations with health outcomes (*28*). This ongoing prospective cohort study enrolled 30,239 participants across the United States between 2003-2007, with intentional oversampling of Black individuals and those from the southeastern Stroke Belt region. The detailed methods and study design has been described in detail elsewhere (*14*, *28*). Briefly, eligible participants had to be non-Hispanic Black or White adults aged ≥45 years. Exclusion criteria included medical conditions preventing long-term participation, active cancer treatment (defined as receipt of chemotherapy or radiation in the previous 2 years), nursing home residency, and inability to communicate in English.

Enrollment required the completion of both a 45-minute telephone questionnaire and an in-home examination visit, where nurses collected physical measurements and blood samples. The study conducted comprehensive baseline assessments covering demographics, medical history, lifestyle factors, and dietary habits.

#### Outcome definitions

Our primary outcome of interest in the REGARDS was incident stroke. Stroke events were identified through semiannual telephone contacts with participants or proxies to ascertain potential strokes or stroke symptoms. All reported events triggered the collection of medical records, including neuroimaging and diagnostic reports, which physicians centrally reviewed to confirm diagnosis, stroke type, and etiology (*28*). Our analysis included all adjudicated stroke cases through April 1, 2019 (*N*=1,321) and a randomly selected subgroup of participants (*N*=1,056), representing a case-cohort design as previously reported (*31*).

Among the randomly selected subcohort (*N*=1,056), we also evaluated secondary outcomes, including heart failure events and incident myocardial infarction, and mortality outcomes, including all-cause mortality, cancer mortality, and cardiovascular death. Each outcome was adjudicated according to standardized protocols (*40–47*). Follow-up data were available through December 31, 2020, for heart failure, and through March 31, 2024, for myocardial infarction. Mortality data were available for cancer and cardiovascular mortality through December 31, 2021 and for all-cause mortality through March 31, 2024. Participants with prevalent conditions at baseline were included, and statistical models were adjusted for relevant baseline conditions (e.g., history of myocardial infarction, coronary bypass surgery, and history of cancer).

Heart failure events include both hospitalizations and deaths, which were adjudicated by experts using a structured form (*48*), based on signs and symptoms of heart failure collected from chart-level data obtained from the hospitals (*49*). During semiannual telephone calls, REGARDS participants were asked if they had been hospitalized since the last call and the reasons for hospitalization. Medical records for all cardiovascular-related conditions were retrieved and reviewed. Adjudication was based on medical records: 1) clinical presentation (symptoms of dyspnea on exertion, paroxysmal nocturnal dyspnea, night cough; and signs including peripheral edema, jugular venous distention, pulmonary rales, hepatomegaly, abnormal central venous pressure, tachycardia), 2) laboratory evaluation (elevated B-type natriuretic peptide), 3) imaging (chest radiogram with cardiomegaly, pulmonary vascular congestion, or pleural effusion; or cardiac imaging such as echocardiography), and 4) clinical response to treatment (weight loss of ≥4.5 kg in 5 days with diuresis).

Myocardial infarction events were defined as (1) a clinical presentation consistent with myocardial ischemia; (2) a rising and/or falling pattern of troponin over at least 6 hours with a peak at least twice the upper limit of normal; or (3) imaging findings consistent with ischemia (*50*).

All-cause mortality was ascertained through semi-annual follow-up calls with participants or proxies (*41*), linkage to the Social Security Death Index and National Death Index, and review of death certificates and medical records. A central adjudication committee reviewed all available data to confirm dates and causes of death. Both proxy-reported death and cause of death ascertained through Social Security Death Index and the National Death Index were found to be highly concordant with expert committee review of medical records and death certificates (*41*).

Cancer mortality was defined as deaths in which cancer was the primary underlying cause, as previously described (*47*, *51*). Time to cancer death was calculated as the date from the baseline REGARDS visit to the date of death listed on death certificate, the Social Security Death Index, the National Death Index, or a published obituary.

Cardiovascular death was defined as death from myocardial infarction, stroke, sudden death, heart failure, or other cardiac causes (*52*). Each death was classified as definite, probable, or possible cardiovascular death based on the strength of the available clinical evidence. The main underlying cause of death was determined by two trained adjudicators who examined all available information, including interviews with next of kin, death certificates, autopsy reports, medical history, and the National Death Index (*50*).

#### Covariate definitions

Age, race, sex, and smoking history were determined by self-report. Body mass index (BMI) was defined as the mass in kilograms divided by the height in meters squared, measured at the in-home visit (*53*). History of cancer was defined based on participant self-report for prior cancer diagnosis. Left ventricular hypertrophy was assessed by standard criteria from the electrocardiogram, and atrial fibrillation was defined by baseline electrocardiogram or self-report for prior atrial fibrillation diagnosis (*29*). Prevalent hypertension was defined by either (1) blood pressure measurement at baseline during the in-person visit, (2) self-reported usage of antihypertensive medications, or (3) a self-reported history of hypertension. The blood pressure was recorded as the average of 2 measurements after the participant had been seated for 5 minutes. A systolic blood pressure ≥130 mmHg or diastolic blood pressure ≥80 mm Hg was used to define hypertension (*54*). Diabetes mellitus was defined as a fasting glucose ≥126 mg/dL (≥7.0 mmol/L), a non-fasting glucose ≥200 mg/dL (≥11.1 mmol/L), or self-reported use of diabetes medications, and coronary artery disease was determined by a self-reported history of myocardial infarction, coronary revascularization procedure, or electrocardiographic evidence of prior myocardial infarction at baseline (*51*). Dyslipidemia was defined as serum total cholesterol level 240 mg/dL, low-density lipoprotein cholesterol level 160 mg/dL, high-density lipoprotein cholesterol level 40 mg/dL, or self-reported medications for dyslipidemia (*34*).

### Ultra-processed food assessments

Based on detailed, itemized cafeteria purchase records from the ChooseWell 365 study, a total of 2,662 unique food item identifiers were categorized into one of four NOVA groups, including unprocessed or minimally processed foods (NOVA 1), processed culinary ingredients (NOVA 2), processed foods (NOVA 3), and UPFs (NOVA 4) (*55*). Three independent reviewers completed the classifications based on published literature and nutritional databases (*32*, *33*). Discrepancies were resolved through consensus discussions, with particular attention to items of ambiguous processing levels. When insufficient information was available to determine processing methods (e.g., sliced meats with unknown details of preservation methods), items were designated with a primary and an alternative classification. Sensitivity analyses were conducted by using the alternative categorizations of ambiguous food items. For each participant, the proportion of UPF purchases was calculated as the percentage of total caloric purchases from NOVA 4 foods relative to total purchases across all NOVA categories throughout the study period.

In the REGARDS cohort, dietary intake was self-reported through a validated Block 1998 Food Frequency Questionnaire (FFQ), and UPF consumption was classified according to the NOVA criteria as previously reported (*14*). Calorie information was sourced from the U.S. Department of Agriculture Food Composition Database and was used to estimate the calories per gram for FFQ items, with UPF intake calculated based on an energy ratio (% of total calories consumed) and total grams of food (% of total grams consumed).

### Metabolomics analysis

Blood samples were collected at the 2-year follow-up visit in the ChooseWell 365 cohort and at the baseline visit in REGARDS. All participants were instructed to fast before venous blood collection. Plasma was separated, aliquoted, and stored at -80°C until metabolomics analysis. Metabolites were extracted using protein precipitation, where 30 μL of K EDTA plasma was first mixed with 30 μL of an internal standard solution containing stable isotope-labeled metabolites (*18*, *31*). Proteins were precipitated using 110 μL of ice-cold acetonitrile: methanol (75:25). After centrifugation, supernatants were analyzed using liquid chromatography-tandem mass spectrometry (LC-MS/MS) (*18*, *31*).

Chromatographic separation was performed using an XBridge Amide column on an Infinity II 1290 high-performance liquid chromatography system with automated alternating column regeneration. Metabolites were detected using an Agilent 6495 triple quadrupole mass spectrometer in dynamic multiple reaction monitoring mode. Both positive and negative ionization modes were employed, with two injections per sample to acquire data in each mode.

Quality control procedures included standardized sample handling protocols with minimal freeze-thaw cycles, system suitability checks at the start of each analytical run, and regular injection of pooled plasma samples for monitoring instrumental performance. Data processing used Agilent MassHunter QQQ Quantitative Analysis software. Peak areas were normalized using a nearest pooled plasma normalization approach, where analyte values were standardized relative to signals from pooled human plasma quality control samples injected every 10 samples.

### Statistical analysis

Metabolomics data were inverse rank-transformed to mitigate the influence of outliers and improve normality. Linear regression models were used to identify associations between UPF consumption based on NOVA1-4 score and circulating metabolites. To account for multiple hypothesis testing, the Bonferroni correction was applied for metabolites that were detected in both cohorts (*N*=160).

Candidate metabolites were first identified in ChooseWell 365 based on their association with UPF consumption derived from food purchasing records in models adjusting for age, sex, race, and smoking status. The robustness of these associations was assessed through sensitivity analyses using alternative food categorizations for ambiguous items. Metabolite-UPF associations were internally validated by the assessment of overall UPF intake derived from 24-hour recall data with the UPF-metabolite index. Food items from the 24-hour recall data were classified according to NOVA criteria by two independent reviewers using the same methodology and reference sources (*32*, *33*) as applied to the cafeteria purchase data.

The UPF-metabolite index was externally validated in the independent REGARDS cohort, where UPF intake was determined from FFQ data. To ensure consistency across different measurement approaches, validation analyses in REGARDS incorporated both the percentage of calories and the percentage of grams-adjusted NOVA values.

To investigate the association between UPF-related metabolites and stroke risk, a composite UPF-metabolite index was constructed by summing standardized (z-scored) metabolite values. For metabolites inversely associated with UPF intake, z-scores were multiplied by −1 to enable a single composite score where higher values indicated greater UPF consumption.

Cox proportional hazard models were used to calculate the hazard ratios (HR). The base models were adjusted for demographics (age at baseline, sex, race, and age by race interaction term), and the fully adjusted models additionally included stroke-related vascular risk factors of smoking status, left ventricular hypertrophy, atrial fibrillation, hypertension, diabetes, and coronary artery disease (*18*, *29*, *56*).

For mediation analysis, UPF consumption was dichotomized at the median to enable the implementation of inverse odds ratio weighting (IORW). The IORW approach was used to assess whether the relationship between UPF consumption and stroke risk was mediated through the UPF-metabolite index (*57*). To evaluate mediation, the UPF-metabolite index was added to the baseline adjusted model. We estimated indirect, direct, and total effects using 1,000 bootstrap replications to obtain bias-corrected confidence intervals. The proportion of the total effect mediated through the UPF-metabolite index was calculated as the ratio of the indirect effect to the total effect. Analyses in the stroke case-cohort sample incorporated survey weights to account for the REGARDS sampling design (*28*).

To examine the relationship between the UPF-metabolite index and additional outcome measures, Cox proportional hazards models for all-cause mortality, cancer mortality, cardiovascular death, heart failure events, and incident myocardial infarction were constructed in the random cohort samples. Base models were adjusted for demographic characteristics (age at baseline, race, and sex). Fully adjusted models included established cardiovascular risk factors previously reported in REGARDS analyses, including smoking status, body mass index, hypertension, diabetes mellitus, and dyslipidemia (*42*, *52*, *58*). To account for key baseline covariates relevant to our outcome assessments, these models also included coronary artery disease, history of myocardial infarction, and history of cancer. Analyses were restricted to the random cohort, to ensure a representative population sample. The proportional hazards assumption was evaluated for all models using Schoenfeld residuals.

For clinical variables and covariates, we employed listwise deletion (complete case analysis) in each model. Information on the extent of missing data for both cohorts is provided in Supplementary Table 2. For the discovery and validation analysis, a Bonferroni-corrected *P* < 3.13 x 10^-4^ was considered significant. For association with health outcomes and in IORW mediation, a *P* < 0.05 was considered significant. Statistical models were constructed in Stata (v.17), and figures were generated in R (v.4.5.0).

### Ethics

All participants included in the study provided written informed consent under study protocols reviewed and approved by the institutional review boards. The ChooseWell 365 study was reviewed and approved by the Mass General Brigham institutional review board. For the REGARDS study, participants provided written informed consent during the in-home examination visit, and the study was conducted under institutional review board approval at the participating institutions. All procedures were in accordance with the ethical standards of the institutional and national research committees, as well as with the 1975 Helsinki Declaration and its later amendments.

## Data Availability

All data are available in the main text or the supplementary materials. The REGARDS metabolomics data for this study have been deposited in the database Brain Data Science Platform (BDSP) https://bdsp.io/content/tmrsccs/1.0.0/ and is available after completion of a data use agreement on the website.
Additional information from the REGARDS study can be requested through the REGARDS study data repository at the University of Alabama at Birmingham: https://www.uab.edu/soph/regardsstudy/.
Metabolomics data from the ChooseWell 365 study are available to qualified researchers after completion of a data use agreement. Requests for this data should be submitted to the corresponding author.

https://www.uab.edu/soph/regardsstudy/

## Acknowledgments

The authors thank the other investigators, the staff, and the participants of the ChooseWell 365 and the REGARDS study for their valuable contributions. A full list of participating REGARDS investigators and institutions can be found at: https://www.uab.edu/soph/regardsstudy/.

## Author contributions

All authors played an instrumental role in the creation of this manuscript. All co-authors critically reviewed the manuscript for important intellectual content. All authors approved the final version of the manuscript.

Conceptualization: WTK, ANT, DEL, ZA

Methodology: WTK, ZA, JD, MRI, HKT

Investigation: ZA, WTK, JD

Purchase item NOVA assignments: ZA, WTK, JD

24hr dietary recall NOVA assignments: ZA, WTK

NOVA assignments in the REGARDS study: CRO

Visualization: ZA

Supervision: WTK

Writing—original draft: ZA, WTK

Writing—review & editing: VMB, JD, AD, ALGG, CRO, SJ, HKT, LCP, MRI, DEL, ANT

## Data and materials availability

All data are available in the main text or the supplementary materials. The REGARDS metabolomics data for this study have been deposited in the database Brain Data Science Platform (BDSP) https://bdsp.io/content/tmrsccs/1.0.0/ and is available after completion of a data use agreement on the website.

Additional information from the REGARDS study can be requested through the REGARDS study data repository at the University of Alabama at Birmingham: https://www.uab.edu/soph/regardsstudy/.

Metabolomics data from the ChooseWell 365 study are available to qualified researchers after completion of a data use agreement. Requests for this data should be submitted to the corresponding author.

## References

1. B. Srour, L. K. Fezeu, E. Kesse-Guyot, B. Allès, C. Méjean, R. M. Andrianasolo, E. Chazelas, M. Deschasaux, S. Hercberg, P. Galan, C. A. Monteiro, C. Julia, M. Touvier, Ultra-processed food intake and risk of cardiovascular disease: prospective cohort study (NutriNet-Santé). BMJ 365, l1451 (2019).

2. N. Gomes Gonçalves, N. Vidal Ferreira, N. Khandpur, E. Martinez Steele, R. Bertazzi Levy, P. Andrade Lotufo, I. M. Bensenor, P. Caramelli, S. M. Alvim de Matos, D. M. Marchioni, C. K. Suemoto, Association Between Consumption of Ultraprocessed Foods and Cognitive Decline. JAMA Neurol, doi: 10.1001/jamaneurol.2022.4397 (2022).

3. H. Li, S. Li, H. Yang, Y. Zhang, S. Zhang, Y. Ma, Y. Hou, X. Zhang, K. Niu, Y. Borné, Y. Wang, Association of Ultraprocessed Food Consumption With Risk of Dementia: A Prospective Cohort Study. Neurology 99, e1056–e1066 (2022).

4. K. Mendoza, S. A. Smith-Warner, S. L. Rossato, N. Khandpur, J. E. Manson, L. Qi, E. B. Rimm, K. J. Mukamal, W. C. Willett, M. Wang, F. B. Hu, J. Mattei, Q. Sun, Ultra-processed foods and cardiovascular disease: analysis of three large US prospective cohorts and a systematic review and meta-analysis of prospective cohort studies. The Lancet Regional Health - Americas 37, 100859 (2024).

5. A. E. Henney, C. S. Gillespie, U. Alam, T. J. Hydes, C. E. Mackay, D. J. Cuthbertson, High intake of ultra-processed food is associated with dementia in adults: a systematic review and meta-analysis of observational studies. J Neurol 271, 198–210 (2024).

6. C. A. Monteiro, G. Cannon, J.-C. Moubarac, R. B. Levy, M. L. C. Louzada, P. C. Jaime, The UN Decade of Nutrition, the NOVA food classification and the trouble with ultra-processing. Public Health Nutr 21, 5–17 (2018).

7. C. A. Monteiro, G. Cannon, R. B. Levy, J.-C. Moubarac, M. L. Louzada, F. Rauber, N. Khandpur, G. Cediel, D. Neri, E. Martinez-Steele, L. G. Baraldi, P. C. Jaime, Ultra-processed foods: what they are and how to identify them. Public Health Nutr 22, 936–941 (2019).

8. G. Menichetti, B. Ravandi, D. Mozaffarian, A.-L. Barabási, Machine learning prediction of the degree of food processing. Nat Commun 14, 2312 (2023).

9. B. Ravandi, G. Ispirova, M. Sebek, P. Mehler, A.-L. Barabási, G. Menichetti, Prevalence of processed foods in major US grocery stores. Nat Food, 1–13 (2025).

10. K. G. Volpp, S. A. Berkowitz, S. V. Sharma, C. A. M. Anderson, L. C. Brewer, M. S. V. Elkind, C. D. Gardner, J. E. Gervis, R. A. Harrington, M. Herrero, A. H. Lichtenstein, M. McClellan, J. Muse, C. A. Roberto, J. P. V. Zachariah, American Heart Association, Food Is Medicine: A Presidential Advisory From the American Heart Association. Circulation 148, 1417–1439 (2023).

11. D. Mozaffarian, J. R. Andrés, E. Cousin, W. H. Frist, D. R. Glickman, The White House Conference on Hunger, Nutrition and Health is an opportunity for transformational change. Nat Food 3, 561–563 (2022).

12. T. Fiolet, B. Srour, L. Sellem, E. Kesse-Guyot, B. Allès, C. Méjean, M. Deschasaux, P. Fassier, P. Latino-Martel, M. Beslay, S. Hercberg, C. Lavalette, C. A. Monteiro, C. Julia, M. Touvier, Consumption of ultra-processed foods and cancer risk: results from NutriNet-Santé prospective cohort. BMJ 360, k322 (2018).

13. Z. Zhang, S. L. Jackson, E. Martinez, C. Gillespie, Q. Yang, Association between ultraprocessed food intake and cardiovascular health in US adults: a cross-sectional analysis of the NHANES 2011-2016. Am J Clin Nutr 113, 428–436 (2021).

14. V. M. Bhave, C. R. Oladele, Z. Ament, N. Kijpaisalratana, A. C. Jones, C. A. Couch, A. Patki, A.-L. Garcia Guarniz, A. Bennett, M. Crowe, M. R. Irvin, W. T. Kimberly, Associations Between Ultra-Processed Food Consumption and Adverse Brain Health Outcomes. Neurology 102, e209432 (2024).

15. V. M. Bhave, Z. Ament, A. Patki, Y. Gao, N. Kijpaisalratana, B. Guo, N. S. Chaudhary, A.-L. G. Guarniz, R. Gerszten, A. Correa, M. Cushman, S. Judd, M. R. Irvin, W. T. Kimberly, Plasma Metabolites Link Dietary Patterns to Stroke Risk. Ann Neurol 93, 500–510 (2023).

16. T. Rafiq, S. M. Azab, K. K. Teo, L. Thabane, S. S. Anand, K. M. Morrison, R. J. de Souza, P. Britz-McKibbin, Nutritional Metabolomics and the Classification of Dietary Biomarker Candidates: A Critical Review. Adv Nutr 12, 2333–2357 (2021).

17. M. Guasch-Ferré, S. N. Bhupathiraju, F. B. Hu, Use of Metabolomics in Improving Assessment of Dietary Intake. Clin Chem 64, 82–98 (2018).

18. Z. Ament, A. Patki, N. Chaudhary, V. M. Bhave, A.-L. Garcia Guarniz, Y. Gao, R. E. Gerszten, A. Correa, S. E. Judd, M. Cushman, D. L. Long, M. R. Irvin, W. T. Kimberly, Nucleosides Associated With Incident Ischemic Stroke in the REGARDS and JHS Cohorts. Neurology 98, e2097–e2107 (2022).

19. D. Vojinovic, M. Kalaoja, S. Trompet, K. Fischer, M. J. Shipley, S. Li, A. S. Havulinna, M. Perola, V. Salomaa, Q. Yang, N. Sattar, P. Jousilahti, N. Amin, C. L. Satizabal, N. Taba, B. Sabayan, R. S. Vasan, M. A. Ikram, D. J. Stott, M. Ala-Korpela, J. W. Jukema, S. Seshadri, J. Kettunen, M. Kivimaki, T. Esko, C. M. van Duijn, Association of Circulating Metabolites in Plasma or Serum and Risk of Stroke: Meta-analysis From 7 Prospective Cohorts. Neurology 96, e1110–e1123 (2021).

20. D. Sun, S. Tiedt, B. Yu, X. Jian, R. F. Gottesman, T. H. Mosley, E. Boerwinkle, M. Dichgans, M. Fornage, A prospective study of serum metabolites and risk of ischemic stroke. Neurology 92, e1890–e1898 (2019).

21. S. Cheng, S. H. Shah, E. J. Corwin, O. Fiehn, R. L. Fitzgerald, R. E. Gerszten, T. Illig, E. P. Rhee, P. R. Srinivas, T. J. Wang, M. Jain, American Heart Association Council on Functional Genomics and Translational Biology; Council on Cardiovascular and Stroke Nursing; Council on Clinical Cardiology; and Stroke Council, Potential Impact and Study Considerations of Metabolomics in Cardiovascular Health and Disease: A Scientific Statement From the American Heart Association. Circ Cardiovasc Genet 10, e000032 (2017).

22. T. J. Wang, M. G. Larson, R. S. Vasan, S. Cheng, E. P. Rhee, E. McCabe, G. D. Lewis, C. S. Fox, P. F. Jacques, C. Fernandez, C. J. O’Donnell, S. A. Carr, V. K. Mootha, J. C. Florez, A. Souza, O. Melander, C. B. Clish, R. E. Gerszten, Metabolite profiles and the risk of developing diabetes. Nat Med 17, 448–453 (2011).

23. Y. Fan, C. Hu, X. Xie, Y. Weng, C. Chen, Z. Wang, X. He, D. Jiang, S. Huang, Z. Hu, F. Liu, Effects of diets on risks of cancer and the mediating role of metabolites. Nat Commun 15, 5903 (2024).

24. A. J. Cross, S. C. Moore, S. Boca, W.-Y. Huang, X. Xiong, R. Stolzenberg-Solomon, R. Sinha, J. N. Sampson, A prospective study of serum metabolites and colorectal cancer risk. Cancer 120, 3049–3057 (2014).

25. D. E. Levy, E. D. Gelsomin, E. B. Rimm, M. Pachucki, J. Sanford, E. Anderson, C. Johnson, R. Schutzberg, A. N. Thorndike, Design of ChooseWell 365: Randomized controlled trial of an automated, personalized worksite intervention to promote healthy food choices and prevent weight gain. Contemporary Clinical Trials 75, 78–86 (2018).

26. A. N. Thorndike, J. L. McCurley, E. D. Gelsomin, E. Anderson, Y. Chang, B. Porneala, C. Johnson, E. B. Rimm, D. E. Levy, Automated Behavioral Workplace Intervention to Prevent Weight Gain and Improve Diet: The ChooseWell 365 Randomized Clinical Trial. JAMA Netw Open 4, e2112528 (2021).

27. V. M. Bhave, Z. Ament, D. E. Levy, A. N. Thorndike, W. T. Kimberly, Workplace food purchases, dietary intake, and gut microbial metabolites in a secondary analysis of the ChooseWell 365 study. Am J Clin Nutr, S0002–9165(24)00444–1 (2024).

28. V. J. Howard, M. Cushman, L. Pulley, C. R. Gomez, R. C. Go, R. J. Prineas, A. Graham, C. S. Moy, G. Howard, The Reasons for Geographic and Racial Differences in Stroke Study: Objectives and Design. Neuroepidemiology 25, 135–143 (2005).

29. Z. Ament, A. Patki, V. M. Bhave, N. S. Chaudhary, A.-L. Garcia Guarniz, N. Kijpaisalratana, S. E. Judd, M. Cushman, D. L. Long, M. R. Irvin, W. T. Kimberly, Gut microbiota-associated metabolites and risk of ischemic stroke in REGARDS. J Cereb Blood Flow Metab 43, 1089–1098 (2023).

30. S. E. Judd, O. M. Gutiérrez, P. Newby, G. Howard, V. J. Howard, J. L. Locher, B. M. Kissela, J. M. Shikany, Dietary patterns are associated with incident stroke and contribute to excess risk of stroke in Black Americans. Stroke 44, 3305–3311 (2013).

31. Z. Ament, N. Kijpaisalratana, V. M. Bhave, C. A. Couch, A.-L. Garcia Guarniz, A. Patki, M. Cushman, S. E. Judd, M. R. Irvin, W. T. Kimberly, Targeted Metabolomics in the REasons for Geographic and Racial Differences in Stroke (REGARDS) Study. Sci Data 12, 395 (2025).

32. E. Martinez-Steele, N. Khandpur, C. Batis, M. Bes-Rastrollo, M. Bonaccio, G. Cediel, I. Huybrechts, F. Juul, R. B. Levy, M. L. da Costa Louzada, P. P. Machado, J.-C. Moubarac, T. Nansel, F. Rauber, B. Srour, M. Touvier, C. A. Monteiro, Best practices for applying the Nova food classification system. Nat Food 4, 445–448 (2023).

33. L. Elizabeth, P. Machado, M. Zinöcker, P. Baker, M. Lawrence, Ultra-Processed Foods and Health Outcomes: A Narrative Review. Nutrients 12, 1955 (2020).

34. D. G. Warnock, P. Muntner, P. A. McCullough, X. Zhang, L. A. McClure, N. Zakai, M. Cushman, B. B. Newsome, R. Kewalramani, M. W. Steffes, G. Howard, W. M. McClellan, Kidney Function, Albuminuria, and All-Cause Mortality in the REGARDS (Reasons for Geographic and Racial Differences in Stroke) Study. American Journal of Kidney Diseases 56, 861–871 (2010).

35. W. McClellan, R. Speckman, L. McClure, V. Howard, R. C. Campbell, M. Cushman, P. Audhya, G. Howard, D. G. Warnock, Prevalence and Characteristics of a Family History of End-Stage Renal Disease among Adults in the United States Population: Reasons for Geographic and Racial Differences in Stroke (REGARDS) Renal Cohort Study. Journal of the American Society of Nephrology 18, 1344–1352 (2007).

36. L. Abar, E. M. Steele, S. K. Lee, L. Kahle, S. C. Moore, E. Watts, C. P. O’Connell, C. E. Matthews, K. A. Herrick, K. D. Hall, L. E. O’Connor, N. D. Freedman, R. Sinha, H. G. Hong, E. Loftfield, Identification and validation of poly-metabolite scores for diets high in ultra-processed food: An observational study and post-hoc randomized controlled crossover-feeding trial. PLoS Med 22, e1004560 (2025).

37. A. L. Brichacek, M. Florkowski, E. Abiona, K. M. Frank, Ultra-Processed Foods: A Narrative Review of the Impact on the Human Gut Microbiome and Variations in Classification Methods. Nutrients 16, 1738 (2024).

38. A. N. Thorndike, J. Riis, L. M. Sonnenberg, D. E. Levy, Traffic-light labels and choice architecture: promoting healthy food choices. Am J Prev Med 46, 143–149 (2014).

39. A. N. Thorndike, L. Sonnenberg, J. Riis, S. Barraclough, D. E. Levy, A 2-phase labeling and choice architecture intervention to improve healthy food and beverage choices. Am J Public Health 102, 527–533 (2012).

40. M. M. Kittleson, K. Breathett, B. Ziaeian, D. Aguilar, V. Blumer, B. Bozkurt, R. L. Diekemper, M. P. Dorsch, P. A. Heidenreich, C. Y. Jurgens, P. Khazanie, G. A. Koromia, H. G. C. Van Spall, 2024 Update to the 2020 ACC/AHA Clinical Performance and Quality Measures for Adults With Heart Failure: A Report of the American Heart Association/American College of Cardiology Joint Committee on Performance Measures. Circ Cardiovasc Qual Outcomes 17, e000132 (2024).

41. J. H. Halanych, F. Shuaib, G. Parmar, R. Tanikella, V. J. Howard, D. L. Roth, R. J. Prineas, M. M. Safford, Agreement on cause of death between proxies, death certificates, and clinician adjudicators in the Reasons for Geographic and Racial Differences in Stroke (REGARDS) study. Am J Epidemiol 173, 1319–1326 (2011).

42. C. P. Walther, O. M. Gutiérrez, M. Cushman, S. E. Judd, J. Lang, W. McClellan, P. Muntner, M. J. Sarnak, M. G. Shlipak, D. G. Warnock, R. Katz, J. H. Ix, Serum albumin concentration and risk of end-stage renal disease: the REGARDS study. Nephrol Dial Transplant 33, 1770–1777 (2018).

43. L. D. Colantonio, E. B. Levitan, H. Yun, M. L. Kilgore, J. D. Rhodes, G. Howard, M. M. Safford, P. Muntner, Use of Medicare claims data for the identification of myocardial infarction: the REasons for Geographic And Racial Differences in Stroke (REGARDS) study. Med Care 56, 1051–1059 (2018).

44. P. Muntner, O. M. Gutiérrez, H. Zhao, C. S. Fox, N. C. Wright, J. R. Curtis, W. McClellan, H. Wang, M. Kilgore, D. G. Warnock, C. B. Bowling, Validation Study of Medicare Claims to Identify Older US Adults With CKD Using the Reasons for Geographic and Racial Differences in Stroke (REGARDS) Study. Am J Kidney Dis 65, 249–258 (2015).

45. O. T. Olubowale, M. M. Safford, T. M. Brown, R. W. Durant, V. J. Howard, C. Gamboa, S. P. Glasser, J. D. Rhodes, E. B. Levitan, Comparison of Expert Adjudicated Coronary Heart Disease and Cardiovascular Disease Mortality With the National Death Index: Results From the REasons for Geographic And Racial Differences in Stroke (REGARDS) Study. J Am Heart Assoc 6, e004966 (2017).

46. H. Bhatt, M. Safford, S. Glasser, Coronary heart disease risk factors and outcomes in the twenty-first century: findings from the REasons for Geographic and Racial Differences in Stroke (REGARDS) Study. Curr Hypertens Rep 17, 541 (2015).

47. J. X. Moore, S. J. Carter, V. Williams, S. Khan, M. W. Lewis-Thames, K. Gilbert, G. Howard, Physical health composite and risk of cancer mortality in the REasons for Geographic and Racial Differences in Stroke Study. Prev Med 132, 105989 (2020).

48. S. R. Heckbert, C. Kooperberg, M. M. Safford, B. M. Psaty, J. Hsia, A. McTiernan, J. M. Gaziano, W. H. Frishman, J. D. Curb, Comparison of self-report, hospital discharge codes, and adjudication of cardiovascular events in the Women’s Health Initiative. Am J Epidemiol 160, 1152–1158 (2004).

49. P. Goyal, M. Mefford, L. Chen, M. Sterling, R. Durant, M. Safford, E. Levitan, Assembling and validating a heart failure-free cohort from the Reasons for Geographic and Racial Differences in Stroke (REGARDS) study. BMC Medical Research Methodology 20, 53 (2020).

50. L. Zhang, E. Reshetnyak, J. B. Ringel, L. C. Pinheiro, A. Carson, D. M. Cummings, R. W. Durant, G. Malla, M. M. Safford, Social Determinants of Health and Cardiovascular Risk among Adults with Diabetes: The Reasons for Geographic and Racial Differences in Stroke (REGARDS) Study. Diabetes Metab J 48, 1073–1083 (2024).

51. L. C. Pinheiro, E. Reshetnyak, M. R. Sterling, E. B. Levitan, M. M. Safford, P. Goyal, Multiple Vulnerabilities to Health Disparities and Incident Heart Failure Hospitalization in the REGARDS Study. Circ Cardiovasc Qual Outcomes 13, e006438 (2020).

52. N. A. Zakai, J. Minnier, M. M. Safford, I. Koh, M. R. Irvin, S. Fazio, M. Cushman, V. J. Howard, N. Pamir, Race-Dependent Association of High-Density Lipoprotein Cholesterol Levels With Incident Coronary Artery Disease. J Am Coll Cardiol 80, 2104–2115 (2022).

53. C. A. Couch, Z. Ament, A. Patki, N. Kijpaisalratana, V. Bhave, A. C. Jones, N. D. Armstrong, M. Cushman, W. T. Kimberly, M. R. Irvin, Sex-Associated Metabolites and Incident Stroke, Incident Coronary Heart Disease, Hypertension, and Chronic Kidney Disease in the REGARDS Cohort. J Am Heart Assoc 13, e032643 (2024).

54. N. Kijpaisalratana, Z. Ament, A. Patki, V. M. Bhave, A.-L. Garcia Guarniz, S. E. Judd, M. Cushman, L. Long, M. R. Irvin, W. T. Kimberly, Association of Circulating Metabolites With Racial Disparities in Hypertension and Stroke in the REGARDS Study. Neurology, 10.1212/WNL.0000000000207264 (2023).

55. V. Braesco, I. Souchon, P. Sauvant, T. Haurogné, M. Maillot, C. Féart, N. Darmon, Ultra-processed foods: how functional is the NOVA system? Eur J Clin Nutr 76, 1245–1253 (2022).

56. Z. Ament, A. Patki, V. M. Bhave, N. Kijpaisalratana, A. C. Jones, C. A. Couch, R. J. Stanton, P. M. Rist, M. Cushman, S. E. Judd, D. L. Long, M. R. Irvin, W. T. Kimberly, Omega-3 Fatty Acids and Risk of Ischemic Stroke in REGARDS. Transl Stroke Res, doi: 10.1007/s12975-024-01256-7 (2024).

57. Q. C. Nguyen, T. L. Osypuk, N. M. Schmidt, M. M. Glymour, E. J. Tchetgen Tchetgen, Practical Guidance for Conducting Mediation Analysis With Multiple Mediators Using Inverse Odds Ratio Weighting. American Journal of Epidemiology 181, 349–356 (2015).

58. M. Cushman, S. E. Judd, V. J. Howard, B. Kissela, O. M. Gutiérrez, N. S. Jenny, A. Ahmed, E. L. Thacker, N. A. Zakai, N-Terminal Pro–B-type Natriuretic Peptide and Stroke Risk: The Reasons for Geographic and Racial Differences in Stroke Cohort. Stroke 45, 1646–1650 (2014).

