## Supplemental Figures and Tables for "A metabolite index of ultra-processed food intake is associated with stroke, cancer mortality, and all-cause mortality"

Zsuzsanna Ament *et al.*

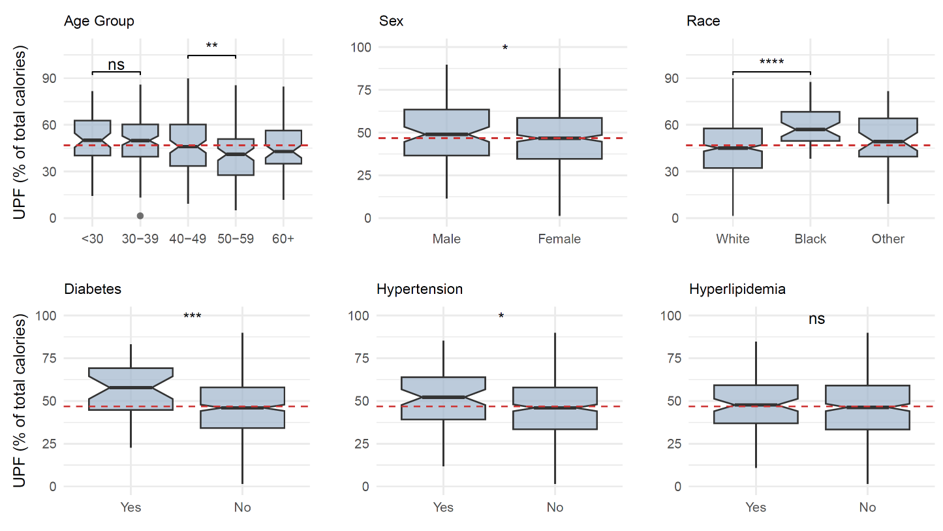

Fig. S1. Overall UPF consumption by demographics and comorbidities in the ChooseWell 365 study. UPF consumption was higher among younger individuals, men, Blacks, and those with diabetes and hypertension. ***, *P* < 0.001; **, *P* < 0.01; *, *P* < 0.05.

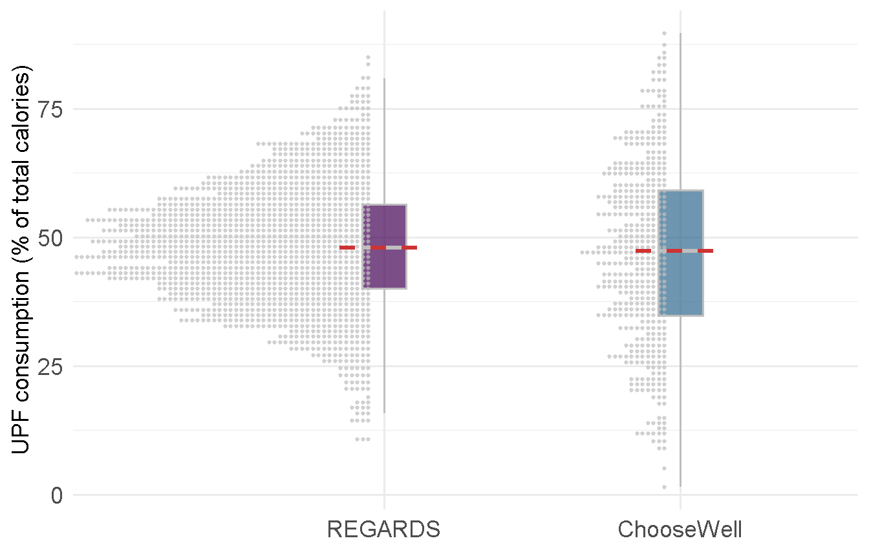

Fig. S2. Distribution of UPF consumption by cohort. The distribution of UPF food consumption among participants in the REGARDS (n=2,165) and Choosewell 365 (n=470) cohorts are shown, as a percentage of total caloric intake. The dashed line is the median and the boxes represent the 25% to 75% interquartile ranges.

**B**

**A**

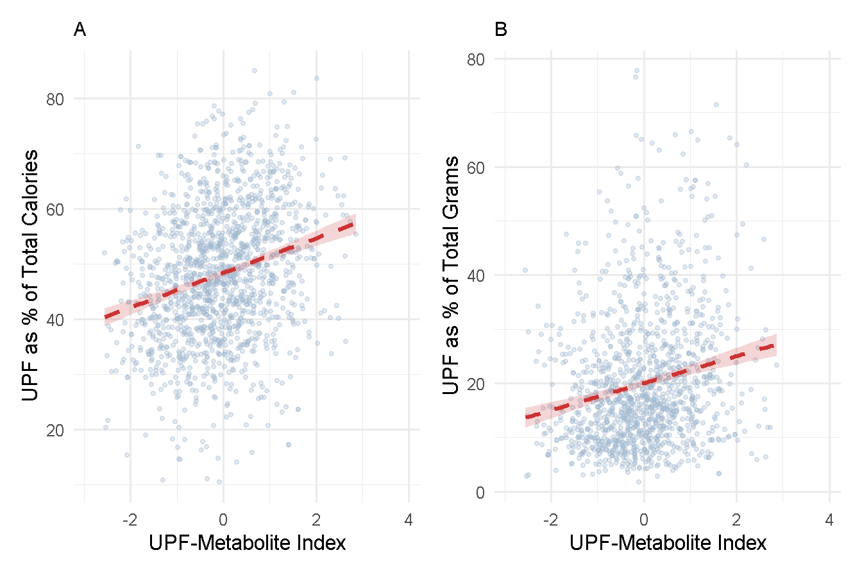

Fig. S3. Associations between the UPF-metabolite index and overall UPF consumption in the REGARDS study. The UPF-metabolite index was associated with UPF intake (A) expressed as a percentage of total calories (*P* < 0.0001), or (B) expressed as a percentage of total grams of intake (*P* < 0.0001).

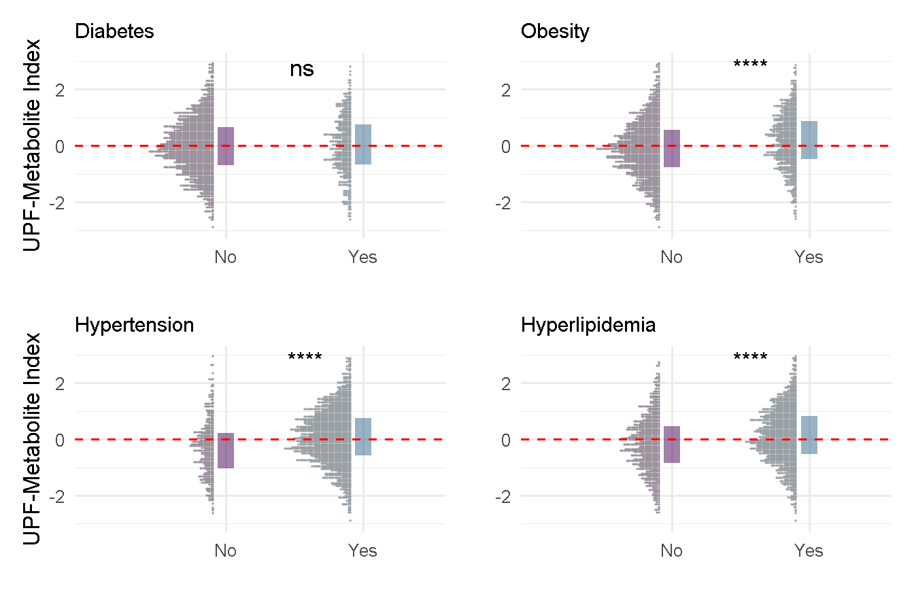

Fig. S4. Distributional differences in the UPF-metabolite index by health status in the REGARDS study. The distribution of the UPF-metabolite index of REGARDS participants with or without diabetes (n.s.), obesity (*P* < 0.0001), hypertension (*P* < 0.0001), and hyperlipidemia (*P* < 0.0001). The dashed line represents the overall median UPF-metabolite index level and the boxes represent the 25% to 75% interquartile range.

Table S1. Individual metabolite associations with overall ultra-processed food (UPF) intake in the ChooseWell 365 and REGARDS study cohorts.

Correlation coefficients and corresponding statistical significance for individual metabolites associated with UPF intake, expressed as the percentage of total calories. In the ChooseWell 365 cohort, models were adjusted for age, race, sex, and smoking status. In the REGARDS cohort, models were adjusted for age, race, sex, smoking status, and the age-by-race interaction term.

This table extends beyond the width of the page and has been provided as a separate pdf file (Table S1.pdf).

Table S2.

Missingness table for the ChooseWell and the REGARDS studies.

|  | **ChooseWell (*N*=470)** | **REGARDS (*N*=2,165)** |
| --- | --- | --- |
| Metabolomics, Missing *N* (%) | 0 (0%) | 0 (0%) |
| UPF consumption (NOVA4) | 0 (0) | 670 (30.9) |
| Age | 0 (0) | 0 (0) |
| Gender/Sex | 0 (0) | 0 (0) |
| Race | 0 (0) | 0 (0) |
| Smoking status | 7 (1.5) | 10 (0.5) |
| Hypertension | 0 (0) | 8 (0.4) |
| Diabetes mellitus | 0 (0) | 25 (1.2) |
| Cardiovascular disease | 5 (1.1) | 34 (1.6) |
| Left ventricular hypertrophy | not assessed | 29 (1.3) |
| Atrial fibrillation | not assessed | 50 (2.3) |
