## Supplemental Table 1 for "A metabolite index of ultra-processed food intake is associated with stroke, cancer mortality, and all-cause mortality"

| | ChooseWell 365 Primary | | | | ChooseWell 365 Sensitivity | | | | REGARDS (% calories adjusted UPFs) | | | | REGARDS (% grams adjusted UPFs) | | | |
|---|---|---|---|---|---|---|---|---|---|---|---|---|---|---|---|---|
| **metabolite** | β | 95%CI | | P | β | 95%CI | | P | β | 95%CI | | P | β | 95%CI | | P |
| glyceric | -0.018 | -0.023 | -0.013 | 4.71E-11 | -0.017 | -0.022 | -0.011 | 1.52E-09 | -0.009 | -0.013 | -0.005 | 2.54E-05 | -0.007 | -0.011 | -0.002 | 3.93E-03 |
| uracil | -0.013 | -0.019 | -0.008 | 1.73E-06 | -0.011 | -0.017 | -0.006 | 7.05E-05 | -0.006 | -0.010 | -0.001 | 9.39E-03 | -0.002 | -0.007 | 0.003 | 4.21E-01 |
| cytidine | 0.013 | 0.007 | 0.018 | 2.18E-06 | 0.011 | 0.006 | 0.016 | 4.30E-05 | 0.006 | 0.002 | 0.011 | 1.86E-03 | 0.007 | 0.002 | 0.011 | 3.61E-03 |
| uridine | -0.013 | -0.019 | -0.008 | 2.31E-06 | -0.012 | -0.017 | -0.006 | 3.90E-05 | -0.010 | -0.014 | -0.006 | 1.01E-06 | -0.007 | -0.012 | -0.003 | 1.36E-03 |
| indole3propanoic | -0.012 | -0.017 | -0.006 | 2.18E-05 | -0.010 | -0.016 | -0.005 | 1.58E-04 | -0.010 | -0.014 | -0.006 | 3.91E-06 | -0.008 | -0.013 | -0.004 | 4.33E-04 |
| ag1deoxyglucose | 0.011 | 0.006 | 0.016 | 4.13E-05 | 0.010 | 0.005 | 0.016 | 1.32E-04 | 0.007 | 0.003 | 0.011 | 9.59E-04 | 0.004 | -0.001 | 0.008 | 1.00E-01 |
| betaisovaleric | -0.011 | -0.017 | -0.006 | 4.66E-05 | -0.011 | -0.016 | -0.005 | 1.21E-04 | -0.008 | -0.013 | -0.004 | 1.02E-04 | -0.004 | -0.009 | 0.000 | 6.96E-02 |
| pseudouridine | 0.011 | 0.006 | 0.016 | 4.91E-05 | 0.010 | 0.005 | 0.015 | 3.15E-04 | 0.003 | -0.001 | 0.007 | 1.67E-01 | 0.006 | 0.002 | 0.010 | 6.73E-03 |
| asparagine | -0.011 | -0.017 | -0.006 | 6.63E-05 | -0.011 | -0.017 | -0.006 | 7.14E-05 | -0.008 | -0.012 | -0.004 | 2.81E-04 | -0.007 | -0.012 | -0.002 | 2.82E-03 |
| guanosine | 0.011 | 0.005 | 0.016 | 7.65E-05 | 0.011 | 0.005 | 0.016 | 1.11E-04 | 0.010 | 0.006 | 0.014 | 5.22E-07 | 0.007 | 0.003 | 0.011 | 1.50E-03 |
| d_gluconic | 0.010 | 0.005 | 0.015 | 1.29E-04 | 0.009 | 0.004 | 0.014 | 4.90E-04 | 0.009 | 0.005 | 0.013 | 2.04E-05 | 0.007 | 0.002 | 0.012 | 3.33E-03 |
| hydroxyglutaric | -0.011 | -0.016 | -0.005 | 1.83E-04 | -0.009 | -0.015 | -0.004 | 1.20E-03 | -0.012 | -0.017 | -0.008 | 7.53E-09 | -0.007 | -0.012 | -0.003 | 2.32E-03 |
| uricacid | 0.009 | 0.004 | 0.014 | 2.24E-04 | 0.010 | 0.005 | 0.015 | 1.06E-04 | 0.002 | -0.002 | 0.006 | 3.85E-01 | 0.004 | -0.001 | 0.008 | 1.15E-01 |
| proline | 0.010 | 0.005 | 0.015 | 3.00E-04 | 0.007 | 0.001 | 0.012 | 1.49E-02 | 0.005 | 0.001 | 0.009 | 2.21E-02 | 0.004 | -0.001 | 0.008 | 9.99E-02 |
| inosine | 0.010 | 0.005 | 0.015 | 3.37E-04 | 0.011 | 0.006 | 0.017 | 6.66E-05 | -0.001 | -0.006 | 0.003 | 5.17E-01 | -0.004 | -0.008 | 0.001 | 1.50E-01 |
| acetoacetic | -0.010 | -0.016 | -0.005 | 3.83E-04 | -0.009 | -0.015 | -0.003 | 1.47E-03 | 0.002 | -0.002 | 0.006 | 2.87E-01 | 0.001 | -0.004 | 0.006 | 6.61E-01 |
| choline | -0.009 | -0.015 | -0.004 | 7.02E-04 | -0.006 | -0.011 | -0.001 | 2.98E-02 | 0.008 | 0.004 | 0.012 | 2.34E-04 | 0.002 | -0.002 | 0.007 | 3.61E-01 |
| glycochenodeoxycholic | 0.009 | 0.004 | 0.015 | 1.28E-03 | 0.009 | 0.003 | 0.014 | 1.90E-03 | 0.000 | -0.004 | 0.004 | 9.82E-01 | -0.001 | -0.006 | 0.003 | 5.27E-01 |
| glutamine | -0.009 | -0.014 | -0.003 | 1.42E-03 | -0.009 | -0.014 | -0.003 | 2.06E-03 | 0.001 | -0.003 | 0.005 | 7.36E-01 | 0.000 | -0.004 | 0.005 | 8.88E-01 |
| pyroglutamic | -0.009 | -0.014 | -0.003 | 1.68E-03 | -0.007 | -0.012 | -0.001 | 1.87E-02 | -0.008 | -0.012 | -0.004 | 2.67E-04 | -0.007 | -0.012 | -0.003 | 2.29E-03 |
| adenosylhomocysteine_sah | 0.009 | 0.003 | 0.014 | 2.01E-03 | 0.006 | 0.001 | 0.011 | 3.19E-02 | 0.007 | 0.003 | 0.011 | 8.07E-04 | 0.004 | -0.001 | 0.008 | 1.29E-01 |
| cotinine | 0.004 | 0.002 | 0.007 | 3.01E-03 | 0.004 | 0.001 | 0.006 | 2.06E-02 | 0.008 | 0.004 | 0.012 | 1.95E-04 | 0.005 | 0.002 | 0.009 | 4.23E-03 |
| xanthine | 0.008 | 0.003 | 0.014 | 3.23E-03 | 0.009 | 0.003 | 0.014 | 1.51E-03 | -0.002 | -0.006 | 0.002 | 3.77E-01 | -0.003 | -0.007 | 0.002 | 2.90E-01 |
| aminoisobutanoic_baiba | -0.008 | -0.013 | -0.002 | 5.96E-03 | -0.005 | -0.010 | 0.001 | 7.96E-02 | -0.002 | -0.006 | 0.002 | 4.17E-01 | 0.002 | -0.003 | 0.006 | 4.60E-01 |
| serine | -0.008 | -0.013 | -0.002 | 6.36E-03 | -0.007 | -0.013 | -0.002 | 9.58E-03 | -0.007 | -0.011 | -0.003 | 5.74E-04 | -0.007 | -0.011 | -0.002 | 3.78E-03 |
| xanthurenic | -0.007 | -0.013 | -0.002 | 7.34E-03 | -0.008 | -0.013 | -0.002 | 4.75E-03 | -0.005 | -0.009 | -0.001 | 2.41E-02 | -0.003 | -0.007 | 0.001 | 1.89E-01 |
| sarcosine | 0.007 | 0.002 | 0.013 | 9.77E-03 | 0.004 | -0.001 | 0.010 | 1.28E-01 | 0.002 | -0.003 | 0.006 | 4.68E-01 | -0.001 | -0.005 | 0.004 | 7.63E-01 |
| indole3carboxylic | 0.007 | 0.002 | 0.013 | 1.06E-02 | 0.006 | 0.000 | 0.012 | 3.31E-02 | 0.004 | 0.000 | 0.008 | 5.23E-02 | 0.004 | 0.000 | 0.009 | 6.77E-02 |
| methyl_cysteine | -0.007 | -0.013 | -0.002 | 1.09E-02 | -0.008 | -0.013 | -0.002 | 5.79E-03 | -0.002 | -0.006 | 0.002 | 3.72E-01 | 0.002 | -0.002 | 0.006 | 3.94E-01 |
| hydroxybutyric3_bhb | -0.007 | -0.013 | -0.002 | 1.13E-02 | -0.005 | -0.011 | 0.000 | 5.74E-02 | -0.006 | -0.010 | -0.002 | 6.99E-03 | -0.001 | -0.005 | 0.004 | 7.38E-01 |
| phosphocreatine_pcr | -0.007 | -0.012 | -0.002 | 1.22E-02 | -0.007 | -0.013 | -0.002 | 8.48E-03 | -0.004 | -0.008 | 0.000 | 5.34E-02 | -0.001 | -0.006 | 0.003 | 5.47E-01 |
| dmgv_dimethylguanidinovaleric | 0.007 | 0.001 | 0.012 | 1.34E-02 | 0.004 | -0.001 | 0.010 | 1.33E-01 | 0.015 | 0.011 | 0.019 | 3.07E-13 | 0.010 | 0.005 | 0.014 | 3.99E-05 |
| allantoin | 0.006 | 0.001 | 0.012 | 1.42E-02 | 0.007 | 0.002 | 0.012 | 1.07E-02 | 0.000 | -0.004 | 0.004 | 9.28E-01 | 0.000 | -0.005 | 0.005 | 9.57E-01 |
| bilirubin | -0.007 | -0.012 | -0.001 | 1.56E-02 | -0.006 | -0.011 | -0.001 | 3.18E-02 | -0.007 | -0.012 | -0.003 | 3.51E-04 | -0.006 | -0.011 | -0.002 | 7.47E-03 |
| anthranilic | 0.007 | 0.001 | 0.012 | 1.68E-02 | 0.007 | 0.002 | 0.013 | 1.12E-02 | -0.001 | -0.005 | 0.003 | 5.91E-01 | -0.001 | -0.005 | 0.004 | 8.23E-01 |
| hypoxanthine | 0.007 | 0.001 | 0.012 | 1.99E-02 | 0.007 | 0.002 | 0.013 | 1.06E-02 | -0.001 | -0.006 | 0.003 | 4.96E-01 | -0.007 | -0.012 | -0.002 | 4.57E-03 |
| acetyl_carnitine | -0.006 | -0.012 | -0.001 | 2.10E-02 | -0.005 | -0.011 | 0.000 | 6.66E-02 | -0.003 | -0.007 | 0.001 | 1.62E-01 | 0.000 | -0.004 | 0.005 | 9.01E-01 |
| arachidonoylglycerolacetate | 0.006 | 0.001 | 0.012 | 2.29E-02 | 0.007 | 0.001 | 0.012 | 1.44E-02 | 0.000 | -0.004 | 0.004 | 8.55E-01 | 0.001 | -0.003 | 0.006 | 6.10E-01 |
| indole3lacticacid | -0.006 | -0.011 | -0.001 | 2.47E-02 | -0.006 | -0.011 | -0.001 | 1.89E-02 | -0.004 | -0.008 | 0.000 | 4.25E-02 | 0.001 | -0.004 | 0.005 | 7.97E-01 |
| malonic | -0.006 | -0.012 | -0.001 | 2.51E-02 | -0.004 | -0.009 | 0.002 | 2.01E-01 | -0.004 | -0.008 | 0.000 | 5.58E-02 | -0.003 | -0.007 | 0.002 | 2.85E-01 |
| alanine | 0.006 | 0.000 | 0.011 | 3.53E-02 | 0.003 | -0.002 | 0.009 | 2.28E-01 | 0.003 | -0.001 | 0.007 | 2.02E-01 | 0.001 | -0.004 | 0.005 | 7.60E-01 |
| fad | -0.006 | -0.011 | 0.000 | 3.63E-02 | -0.007 | -0.012 | -0.001 | 1.54E-02 | -0.002 | -0.006 | 0.002 | 3.40E-01 | -0.001 | -0.005 | 0.004 | 7.79E-01 |
| gssg | -0.006 | -0.011 | 0.000 | 3.93E-02 | -0.007 | -0.013 | -0.002 | 1.10E-02 | 0.001 | -0.003 | 0.005 | 6.84E-01 | 0.000 | -0.005 | 0.005 | 9.84E-01 |
| glycocholic | 0.006 | 0.000 | 0.011 | 4.24E-02 | 0.006 | 0.000 | 0.011 | 4.74E-02 | -0.001 | -0.005 | 0.003 | 7.27E-01 | -0.001 | -0.005 | 0.004 | 7.47E-01 |
| lactic | 0.006 | 0.000 | 0.011 | 4.28E-02 | 0.003 | -0.003 | 0.008 | 3.17E-01 | 0.001 | -0.003 | 0.006 | 5.78E-01 | -0.005 | -0.009 | 0.000 | 5.98E-02 |
| trimethyaminenoxide_tmao | -0.005 | -0.011 | 0.000 | 4.93E-02 | -0.007 | -0.012 | -0.001 | 1.42E-02 | 0.000 | -0.004 | 0.004 | 9.14E-01 | 0.001 | -0.004 | 0.005 | 8.26E-01 |
| orotic | 0.005 | 0.000 | 0.011 | 5.46E-02 | 0.006 | 0.000 | 0.011 | 4.70E-02 | 0.002 | -0.003 | 0.006 | 4.31E-01 | 0.000 | -0.004 | 0.005 | 8.81E-01 |
| cystamine | -0.005 | -0.011 | 0.000 | 5.70E-02 | -0.006 | -0.012 | -0.001 | 2.43E-02 | -0.004 | -0.008 | 0.001 | 8.46E-02 | 0.000 | -0.005 | 0.004 | 8.51E-01 |
| indoxylsulfate | 0.005 | 0.000 | 0.011 | 5.84E-02 | 0.006 | 0.000 | 0.011 | 4.58E-02 | 0.006 | 0.002 | 0.010 | 5.37E-03 | 0.005 | 0.001 | 0.010 | 2.94E-02 |
| hippuric | -0.005 | -0.011 | 0.000 | 5.88E-02 | -0.007 | -0.013 | -0.002 | 1.01E-02 | -0.007 | -0.011 | -0.003 | 1.32E-03 | -0.009 | -0.014 | -0.005 | 5.04E-05 |
| homocysteine | -0.005 | -0.011 | 0.000 | 6.40E-02 | -0.006 | -0.012 | -0.001 | 3.04E-02 | -0.001 | -0.005 | 0.003 | 7.26E-01 | 0.003 | -0.002 | 0.007 | 2.17E-01 |
| glutamate | 0.005 | 0.000 | 0.011 | 6.54E-02 | 0.002 | -0.003 | 0.008 | 4.52E-01 | -0.001 | -0.005 | 0.004 | 8.05E-01 | 0.000 | -0.005 | 0.005 | 9.96E-01 |
| dimethylglycine | 0.005 | 0.000 | 0.011 | 6.58E-02 | 0.003 | -0.002 | 0.009 | 2.59E-01 | 0.004 | 0.000 | 0.008 | 7.39E-02 | 0.001 | -0.003 | 0.005 | 6.54E-01 |
| kynurenine | 0.005 | 0.000 | 0.010 | 7.23E-02 | 0.006 | 0.001 | 0.012 | 2.55E-02 | 0.004 | 0.000 | 0.008 | 3.45E-02 | 0.005 | 0.001 | 0.010 | 2.17E-02 |
| tryptophan | -0.005 | -0.010 | 0.000 | 7.58E-02 | -0.004 | -0.009 | 0.001 | 1.13E-01 | -0.008 | -0.012 | -0.004 | 2.05E-04 | -0.006 | -0.011 | -0.002 | 5.81E-03 |
| cyclicamp | 0.005 | -0.001 | 0.011 | 7.91E-02 | 0.005 | 0.000 | 0.011 | 7.21E-02 | 0.005 | 0.001 | 0.010 | 9.62E-03 | 0.004 | -0.001 | 0.009 | 8.65E-02 |
| hydroxylysine | -0.005 | -0.010 | 0.001 | 9.56E-02 | -0.005 | -0.011 | 0.000 | 7.23E-02 | 0.002 | -0.002 | 0.007 | 2.82E-01 | 0.005 | 0.000 | 0.009 | 4.49E-02 |
| oxalic | -0.005 | -0.010 | 0.001 | 1.01E-01 | -0.003 | -0.008 | 0.003 | 3.70E-01 | -0.006 | -0.011 | -0.002 | 2.30E-03 | -0.006 | -0.010 | -0.001 | 1.33E-02 |
| monomethylarginine_nmma | -0.004 | -0.010 | 0.001 | 1.06E-01 | -0.004 | -0.009 | 0.001 | 1.40E-01 | -0.003 | -0.007 | 0.001 | 1.40E-01 | -0.003 | -0.008 | 0.001 | 1.62E-01 |
| methyl_histidine | -0.004 | -0.010 | 0.001 | 1.14E-01 | -0.002 | -0.008 | 0.003 | 4.20E-01 | -0.003 | -0.008 | 0.001 | 1.19E-01 | -0.001 | -0.006 | 0.004 | 6.37E-01 |
| glucose_fruct_galact | 0.004 | -0.001 | 0.010 | 1.26E-01 | 0.004 | -0.001 | 0.010 | 1.36E-01 | 0.000 | -0.004 | 0.005 | 8.27E-01 | 0.003 | -0.002 | 0.008 | 2.06E-01 |
| taurine | -0.004 | -0.010 | 0.001 | 1.29E-01 | -0.002 | -0.008 | 0.003 | 3.78E-01 | 0.001 | -0.004 | 0.005 | 7.35E-01 | 0.006 | 0.002 | 0.010 | 8.19E-03 |
| c182carnitine | 0.004 | -0.001 | 0.010 | 1.32E-01 | 0.005 | -0.001 | 0.010 | 9.83E-02 | 0.001 | -0.003 | 0.005 | 7.54E-01 | -0.006 | -0.010 | -0.001 | 1.48E-02 |
| valine | -0.004 | -0.010 | 0.001 | 1.37E-01 | -0.005 | -0.010 | 0.001 | 7.52E-02 | 0.000 | -0.005 | 0.004 | 8.28E-01 | 0.006 | 0.002 | 0.010 | 5.61E-03 |
| acetyl_lysineepsilon | 0.004 | -0.001 | 0.010 | 1.38E-01 | 0.004 | -0.001 | 0.010 | 1.49E-01 | 0.001 | -0.003 | 0.005 | 6.16E-01 | 0.005 | 0.001 | 0.010 | 2.30E-02 |
| deoxyuridine2 | 0.004 | -0.001 | 0.010 | 1.40E-01 | 0.004 | -0.002 | 0.009 | 2.04E-01 | -0.004 | -0.008 | 0.000 | 5.51E-02 | -0.005 | -0.010 | 0.000 | 3.63E-02 |
| amp | 0.004 | -0.001 | 0.010 | 1.41E-01 | 0.004 | -0.001 | 0.010 | 1.17E-01 | 0.000 | -0.004 | 0.004 | 9.49E-01 | 0.002 | -0.003 | 0.006 | 4.58E-01 |
| c4methylmalonylcarnitine | 0.004 | -0.002 | 0.009 | 1.79E-01 | 0.004 | -0.002 | 0.009 | 2.08E-01 | -0.004 | -0.009 | 0.000 | 4.47E-02 | 0.004 | 0.000 | 0.009 | 6.43E-02 |
| xanthosine | 0.004 | -0.002 | 0.009 | 1.84E-01 | 0.005 | -0.001 | 0.010 | 9.65E-02 | 0.003 | -0.001 | 0.007 | 1.37E-01 | 0.005 | 0.001 | 0.010 | 2.48E-02 |
| c16carnitine | 0.004 | -0.002 | 0.009 | 1.86E-01 | 0.005 | -0.001 | 0.010 | 1.02E-01 | -0.004 | -0.008 | 0.000 | 5.07E-02 | -0.005 | -0.009 | 0.000 | 4.49E-02 |
| c7carnitine | 0.004 | -0.002 | 0.009 | 1.99E-01 | 0.004 | -0.002 | 0.010 | 1.56E-01 | 0.004 | 0.000 | 0.008 | 4.80E-02 | 0.002 | -0.002 | 0.007 | 3.49E-01 |
| adenosylmethyonine_sam | 0.004 | -0.002 | 0.009 | 2.02E-01 | 0.003 | -0.002 | 0.009 | 2.37E-01 | 0.005 | 0.001 | 0.009 | 2.22E-02 | 0.007 | 0.002 | 0.011 | 4.50E-03 |
| acetyl_neuraminic | 0.003 | -0.002 | 0.009 | 2.03E-01 | 0.004 | -0.001 | 0.010 | 1.02E-01 | 0.007 | 0.003 | 0.011 | 3.63E-04 | 0.007 | 0.003 | 0.012 | 1.06E-03 |
| leucine | -0.003 | -0.009 | 0.002 | 2.05E-01 | -0.005 | -0.010 | 0.001 | 8.06E-02 | 0.003 | -0.001 | 0.007 | 1.42E-01 | 0.004 | -0.001 | 0.008 | 8.53E-02 |
| warfarin | -0.001 | -0.002 | 0.000 | 2.07E-01 | -0.001 | -0.002 | 0.000 | 2.11E-01 | -0.003 | -0.006 | 0.000 | 5.10E-02 | -0.002 | -0.005 | 0.001 | 2.89E-01 |
| pyruvic | -0.004 | -0.009 | 0.002 | 2.08E-01 | -0.005 | -0.010 | 0.001 | 1.07E-01 | 0.004 | 0.000 | 0.008 | 7.86E-02 | -0.001 | -0.006 | 0.003 | 5.98E-01 |
| ketoisovaleric_kiv | -0.003 | -0.009 | 0.002 | 2.13E-01 | -0.004 | -0.009 | 0.002 | 1.78E-01 | -0.007 | -0.011 | -0.003 | 1.59E-03 | -0.004 | -0.008 | 0.000 | 7.39E-02 |
| c162carnitine | 0.004 | -0.002 | 0.009 | 2.15E-01 | 0.004 | -0.002 | 0.009 | 1.70E-01 | 0.002 | -0.002 | 0.006 | 3.63E-01 | -0.001 | -0.006 | 0.003 | 6.44E-01 |
| tyrosine | 0.003 | -0.002 | 0.009 | 2.18E-01 | 0.003 | -0.003 | 0.008 | 3.43E-01 | 0.002 | -0.002 | 0.006 | 3.21E-01 | 0.006 | 0.001 | 0.010 | 1.45E-02 |
| maleic | -0.003 | -0.009 | 0.002 | 2.21E-01 | -0.003 | -0.009 | 0.002 | 2.48E-01 | -0.003 | -0.008 | 0.001 | 1.13E-01 | -0.004 | -0.009 | 0.001 | 8.95E-02 |
| c161carnitine | 0.003 | -0.002 | 0.009 | 2.24E-01 | 0.004 | -0.001 | 0.010 | 1.30E-01 | -0.002 | -0.006 | 0.002 | 2.28E-01 | -0.004 | -0.009 | 0.000 | 6.06E-02 |
| betaalanine | 0.003 | -0.002 | 0.009 | 2.39E-01 | 0.003 | -0.002 | 0.009 | 2.25E-01 | 0.001 | -0.003 | 0.005 | 7.46E-01 | 0.002 | -0.002 | 0.006 | 3.31E-01 |
| taurodeoxycholic | 0.003 | -0.002 | 0.009 | 2.39E-01 | 0.004 | -0.001 | 0.010 | 1.39E-01 | -0.003 | -0.007 | 0.001 | 2.08E-01 | -0.002 | -0.007 | 0.002 | 3.43E-01 |
| c121carnitine | 0.003 | -0.002 | 0.009 | 2.49E-01 | 0.004 | -0.002 | 0.010 | 1.58E-01 | 0.003 | -0.001 | 0.007 | 1.82E-01 | 0.001 | -0.004 | 0.005 | 7.75E-01 |
| c142carnitine | 0.003 | -0.002 | 0.009 | 2.65E-01 | 0.004 | -0.002 | 0.009 | 1.74E-01 | 0.003 | -0.001 | 0.007 | 1.85E-01 | -0.001 | -0.005 | 0.004 | 7.15E-01 |
| c14carnitine | 0.003 | -0.002 | 0.009 | 2.77E-01 | 0.003 | -0.002 | 0.009 | 2.79E-01 | -0.002 | -0.006 | 0.002 | 3.28E-01 | -0.002 | -0.006 | 0.003 | 4.25E-01 |
| ketoisocaproic_kic_ket | -0.003 | -0.008 | 0.002 | 2.77E-01 | -0.002 | -0.007 | 0.004 | 5.64E-01 | -0.008 | -0.012 | -0.004 | 4.25E-05 | -0.007 | -0.011 | -0.003 | 1.02E-03 |
| anserine | 0.003 | -0.003 | 0.008 | 3.03E-01 | 0.002 | -0.003 | 0.008 | 4.54E-01 | 0.004 | 0.001 | 0.008 | 2.58E-02 | 0.003 | -0.001 | 0.008 | 1.16E-01 |
| guanosinemonophosphate_gmp | 0.003 | -0.003 | 0.008 | 3.05E-01 | 0.003 | -0.003 | 0.008 | 3.16E-01 | -0.003 | -0.007 | 0.001 | 1.57E-01 | 0.001 | -0.004 | 0.006 | 6.26E-01 |
| histidine | -0.003 | -0.008 | 0.003 | 3.21E-01 | -0.005 | -0.010 | 0.001 | 1.03E-01 | -0.003 | -0.007 | 0.001 | 2.03E-01 | 0.000 | -0.005 | 0.004 | 9.18E-01 |
| atp | 0.003 | -0.003 | 0.008 | 3.21E-01 | 0.004 | -0.001 | 0.010 | 1.32E-01 | 0.002 | -0.003 | 0.006 | 4.55E-01 | 0.003 | -0.001 | 0.008 | 1.46E-01 |
| deoxycholic | 0.003 | -0.003 | 0.008 | 3.29E-01 | 0.002 | -0.003 | 0.008 | 4.08E-01 | 0.003 | -0.002 | 0.007 | 2.29E-01 | 0.001 | -0.004 | 0.005 | 8.01E-01 |
| c3carnitine | 0.003 | -0.003 | 0.008 | 3.45E-01 | 0.002 | -0.003 | 0.008 | 3.91E-01 | -0.001 | -0.005 | 0.003 | 5.61E-01 | 0.002 | -0.003 | 0.007 | 3.65E-01 |
| hydroxybutyric2 | 0.003 | -0.003 | 0.008 | 3.52E-01 | 0.003 | -0.003 | 0.008 | 2.91E-01 | -0.007 | -0.011 | -0.003 | 9.81E-04 | -0.003 | -0.008 | 0.001 | 1.70E-01 |
| acetyl_glutamine | 0.003 | -0.003 | 0.008 | 3.53E-01 | 0.002 | -0.004 | 0.007 | 5.40E-01 | 0.000 | -0.004 | 0.004 | 9.45E-01 | 0.001 | -0.003 | 0.006 | 6.22E-01 |
| c3malonylcarnitine | 0.003 | -0.003 | 0.008 | 3.56E-01 | 0.003 | -0.002 | 0.009 | 2.25E-01 | 0.001 | -0.003 | 0.006 | 4.88E-01 | 0.001 | -0.004 | 0.005 | 7.16E-01 |
| kynurenic | -0.002 | -0.008 | 0.003 | 3.61E-01 | -0.003 | -0.008 | 0.003 | 3.43E-01 | -0.001 | -0.005 | 0.003 | 4.89E-01 | -0.001 | -0.005 | 0.004 | 7.03E-01 |
| isoleucine | 0.002 | -0.003 | 0.008 | 3.61E-01 | 0.003 | -0.002 | 0.008 | 2.47E-01 | -0.002 | -0.006 | 0.002 | 4.09E-01 | -0.001 | -0.005 | 0.004 | 8.08E-01 |
| carnitine | 0.003 | -0.003 | 0.008 | 3.63E-01 | 0.003 | -0.002 | 0.009 | 2.54E-01 | -0.002 | -0.006 | 0.002 | 4.03E-01 | -0.002 | -0.007 | 0.003 | 3.68E-01 |

| | | | | | | | | | | | | | | | | |
|---|---|---|---|---|---|---|---|---|---|---|---|---|---|---|---|---|
| pantothenic | -0.002 | -0.008 | 0.003 | 3.68E-01 | -0.001 | -0.006 | 0.004 | 7.10E-01 | -0.009 | -0.013 | -0.005 | 9.25E-06 | -0.010 | -0.014 | -0.006 | 7.19E-06 |
| glycerol3phosphate | 0.002 | -0.003 | 0.008 | 3.77E-01 | 0.002 | -0.003 | 0.008 | 4.20E-01 | 0.001 | -0.003 | 0.006 | 5.11E-01 | 0.001 | -0.004 | 0.005 | 7.56E-01 |
| adma_sdma | 0.002 | -0.003 | 0.007 | 3.89E-01 | 0.002 | -0.003 | 0.007 | 4.25E-01 | 0.003 | -0.001 | 0.007 | 1.55E-01 | 0.006 | 0.001 | 0.011 | 9.38E-03 |
| succinic | 0.002 | -0.003 | 0.008 | 3.91E-01 | 0.003 | -0.003 | 0.008 | 3.54E-01 | -0.002 | -0.006 | 0.002 | 4.37E-01 | -0.005 | -0.010 | 0.000 | 3.26E-02 |
| c5valerylcarnitine | 0.002 | -0.003 | 0.008 | 4.12E-01 | 0.003 | -0.002 | 0.009 | 2.70E-01 | 0.001 | -0.003 | 0.005 | 6.54E-01 | 0.003 | -0.002 | 0.007 | 2.56E-01 |
| methionine | -0.002 | -0.008 | 0.003 | 4.16E-01 | -0.003 | -0.009 | 0.002 | 2.74E-01 | 0.001 | -0.003 | 0.005 | 7.77E-01 | -0.002 | -0.007 | 0.002 | 3.27E-01 |
| hydroxyphenylpyruvic | -0.002 | -0.008 | 0.003 | 4.41E-01 | -0.004 | -0.009 | 0.002 | 1.67E-01 | 0.000 | -0.004 | 0.004 | 9.00E-01 | 0.006 | 0.002 | 0.011 | 8.29E-03 |
| c9carnitine | 0.002 | -0.003 | 0.008 | 4.42E-01 | 0.002 | -0.004 | 0.008 | 4.69E-01 | 0.000 | -0.004 | 0.004 | 8.36E-01 | 0.001 | -0.004 | 0.005 | 7.07E-01 |
| phosphoethanolamine | -0.002 | -0.008 | 0.003 | 4.55E-01 | -0.002 | -0.008 | 0.004 | 4.67E-01 | -0.009 | -0.013 | -0.005 | 1.41E-05 | -0.004 | -0.009 | 0.001 | 9.33E-02 |
| thyroxine | -0.002 | -0.008 | 0.003 | 4.69E-01 | -0.002 | -0.007 | 0.004 | 5.78E-01 | 0.004 | 0.000 | 0.009 | 4.13E-02 | -0.002 | -0.007 | 0.003 | 4.11E-01 |
| c6carnitine | 0.002 | -0.004 | 0.008 | 4.80E-01 | 0.003 | -0.003 | 0.008 | 3.72E-01 | 0.001 | -0.003 | 0.005 | 6.39E-01 | -0.003 | -0.008 | 0.001 | 1.52E-01 |
| c181carnitine | 0.002 | -0.004 | 0.007 | 4.86E-01 | 0.003 | -0.003 | 0.008 | 3.53E-01 | -0.001 | -0.005 | 0.003 | 7.18E-01 | -0.006 | -0.010 | -0.001 | 1.38E-02 |
| oxaloacetic | -0.002 | -0.007 | 0.004 | 5.00E-01 | 0.000 | -0.005 | 0.005 | 9.74E-01 | 0.001 | -0.003 | 0.005 | 5.08E-01 | -0.002 | -0.006 | 0.003 | 4.39E-01 |
| adp | 0.002 | -0.004 | 0.007 | 5.25E-01 | 0.003 | -0.003 | 0.008 | 3.63E-01 | -0.002 | -0.006 | 0.002 | 3.26E-01 | 0.000 | -0.005 | 0.005 | 9.58E-01 |
| salicylic | -0.002 | -0.007 | 0.004 | 5.35E-01 | -0.001 | -0.006 | 0.005 | 8.10E-01 | -0.004 | -0.008 | 0.001 | 9.39E-02 | -0.002 | -0.007 | 0.002 | 3.07E-01 |
| c141carnitine | 0.002 | -0.004 | 0.007 | 5.54E-01 | 0.002 | -0.003 | 0.008 | 4.25E-01 | 0.000 | -0.004 | 0.004 | 9.78E-01 | -0.001 | -0.006 | 0.003 | 5.63E-01 |
| udp_glucosegalactose | 0.002 | -0.004 | 0.007 | 5.74E-01 | 0.001 | -0.005 | 0.007 | 7.35E-01 | 0.002 | -0.002 | 0.007 | 2.57E-01 | 0.006 | 0.001 | 0.010 | 2.15E-02 |
| aminoadipic | 0.002 | -0.004 | 0.007 | 5.81E-01 | 0.000 | -0.005 | 0.006 | 9.42E-01 | -0.003 | -0.007 | 0.001 | 1.15E-01 | -0.002 | -0.006 | 0.003 | 4.71E-01 |
| cystathionine | 0.001 | -0.004 | 0.007 | 5.88E-01 | 0.001 | -0.005 | 0.006 | 8.32E-01 | 0.003 | -0.001 | 0.007 | 1.26E-01 | 0.003 | -0.002 | 0.008 | 1.91E-01 |
| anandamide | 0.001 | -0.004 | 0.007 | 6.03E-01 | 0.000 | -0.006 | 0.005 | 9.58E-01 | 0.001 | -0.003 | 0.006 | 5.21E-01 | 0.001 | -0.004 | 0.005 | 7.08E-01 |
| c12carnitine | 0.001 | -0.004 | 0.007 | 6.08E-01 | 0.002 | -0.004 | 0.008 | 4.98E-01 | 0.001 | -0.003 | 0.005 | 7.47E-01 | -0.001 | -0.006 | 0.003 | 6.48E-01 |
| citric_isocitric | 0.001 | -0.004 | 0.007 | 6.10E-01 | 0.002 | -0.003 | 0.007 | 4.48E-01 | -0.002 | -0.006 | 0.002 | 2.55E-01 | -0.001 | -0.005 | 0.004 | 7.39E-01 |
| citidinemonophosphate_cmp | 0.001 | -0.004 | 0.007 | 6.25E-01 | 0.002 | -0.004 | 0.007 | 5.94E-01 | 0.005 | 0.001 | 0.010 | 1.38E-02 | 0.003 | -0.002 | 0.008 | 2.58E-01 |
| guanine | -0.001 | -0.007 | 0.004 | 6.36E-01 | -0.003 | -0.008 | 0.003 | 3.56E-01 | 0.002 | -0.002 | 0.006 | 3.60E-01 | -0.001 | -0.006 | 0.003 | 5.41E-01 |
| acetyl_lysinealpha | -0.001 | -0.007 | 0.004 | 6.43E-01 | -0.001 | -0.007 | 0.004 | 5.92E-01 | 0.002 | -0.002 | 0.006 | 3.48E-01 | 0.005 | 0.000 | 0.009 | 5.54E-02 |
| taurocholic | -0.001 | -0.007 | 0.004 | 6.48E-01 | -0.001 | -0.007 | 0.004 | 7.11E-01 | -0.002 | -0.006 | 0.002 | 3.24E-01 | -0.001 | -0.006 | 0.004 | 6.34E-01 |
| homogentisic | -0.001 | -0.007 | 0.004 | 6.53E-01 | 0.000 | -0.006 | 0.006 | 9.87E-01 | -0.004 | -0.008 | 0.000 | 4.49E-02 | 0.001 | -0.004 | 0.005 | 7.99E-01 |
| c8carnitine | 0.001 | -0.004 | 0.007 | 6.87E-01 | 0.002 | -0.004 | 0.007 | 5.81E-01 | -0.001 | -0.005 | 0.003 | 7.40E-01 | -0.006 | -0.010 | -0.001 | 1.51E-02 |
| c18carnitine | 0.001 | -0.004 | 0.007 | 7.10E-01 | 0.002 | -0.004 | 0.007 | 5.10E-01 | 0.000 | -0.004 | 0.005 | 8.39E-01 | -0.003 | -0.008 | 0.001 | 1.52E-01 |
| glycine | -0.001 | -0.006 | 0.004 | 7.29E-01 | -0.001 | -0.007 | 0.004 | 5.99E-01 | -0.001 | -0.005 | 0.004 | 7.83E-01 | -0.004 | -0.009 | 0.000 | 5.37E-02 |
| udp_glcnac | -0.001 | -0.006 | 0.005 | 7.37E-01 | 0.000 | -0.006 | 0.005 | 9.22E-01 | 0.001 | -0.003 | 0.005 | 6.58E-01 | 0.002 | -0.002 | 0.007 | 3.48E-01 |
| betaine | 0.001 | -0.005 | 0.006 | 7.60E-01 | 0.001 | -0.005 | 0.007 | 7.21E-01 | 0.007 | 0.003 | 0.012 | 5.77E-04 | 0.007 | 0.003 | 0.012 | 1.59E-03 |
| gsh | 0.001 | -0.005 | 0.006 | 7.62E-01 | 0.000 | -0.006 | 0.005 | 9.21E-01 | 0.002 | -0.002 | 0.006 | 3.49E-01 | 0.002 | -0.003 | 0.006 | 4.94E-01 |
| hydroxytyrosine | 0.001 | -0.005 | 0.006 | 7.62E-01 | 0.001 | -0.005 | 0.006 | 8.38E-01 | 0.006 | 0.002 | 0.011 | 3.31E-03 | 0.011 | 0.006 | 0.015 | 1.58E-06 |
| cystine | -0.001 | -0.006 | 0.005 | 7.80E-01 | -0.001 | -0.007 | 0.004 | 6.18E-01 | -0.001 | -0.005 | 0.003 | 6.68E-01 | 0.000 | -0.005 | 0.004 | 8.38E-01 |
| threonine | 0.001 | -0.005 | 0.006 | 7.92E-01 | 0.000 | -0.005 | 0.006 | 9.66E-01 | -0.001 | -0.006 | 0.003 | 4.94E-01 | -0.003 | -0.008 | 0.001 | 1.77E-01 |
| c26carnitine | 0.001 | -0.005 | 0.006 | 7.94E-01 | 0.001 | -0.005 | 0.006 | 7.84E-01 | -0.004 | -0.008 | 0.000 | 4.26E-02 | -0.001 | -0.006 | 0.003 | 5.61E-01 |
| c10carnitine | 0.001 | -0.005 | 0.006 | 8.14E-01 | 0.001 | -0.004 | 0.007 | 6.39E-01 | -0.002 | -0.006 | 0.002 | 3.34E-01 | -0.005 | -0.010 | -0.001 | 2.86E-02 |
| a_ketoglutaric | 0.001 | -0.005 | 0.006 | 8.21E-01 | -0.001 | -0.006 | 0.005 | 7.31E-01 | 0.003 | -0.001 | 0.007 | 1.23E-01 | 0.002 | -0.003 | 0.006 | 5.16E-01 |
| folate | 0.001 | -0.005 | 0.006 | 8.23E-01 | 0.000 | -0.005 | 0.006 | 8.80E-01 | 0.003 | -0.001 | 0.007 | 1.52E-01 | 0.002 | -0.002 | 0.007 | 3.03E-01 |
| arginine | 0.000 | -0.006 | 0.005 | 8.61E-01 | 0.000 | -0.006 | 0.005 | 9.37E-01 | -0.003 | -0.007 | 0.001 | 1.64E-01 | -0.001 | -0.005 | 0.004 | 8.17E-01 |
| aminooctanoate2 | 0.000 | -0.006 | 0.005 | 8.63E-01 | -0.001 | -0.007 | 0.005 | 7.24E-01 | -0.001 | -0.006 | 0.003 | 4.85E-01 | -0.003 | -0.008 | 0.001 | 1.79E-01 |
| fumaric | 0.000 | -0.005 | 0.006 | 8.77E-01 | 0.001 | -0.005 | 0.006 | 8.42E-01 | -0.002 | -0.006 | 0.003 | 4.67E-01 | -0.003 | -0.007 | 0.002 | 2.58E-01 |
| acetyl_glutamate | 0.000 | -0.005 | 0.006 | 8.77E-01 | 0.000 | -0.006 | 0.005 | 9.50E-01 | 0.004 | 0.000 | 0.008 | 6.31E-02 | 0.006 | 0.001 | 0.011 | 1.08E-02 |
| phenylalanine | 0.000 | -0.006 | 0.005 | 8.81E-01 | -0.001 | -0.007 | 0.004 | 6.79E-01 | 0.002 | -0.002 | 0.006 | 3.87E-01 | -0.001 | -0.006 | 0.004 | 6.90E-01 |
| a_glycerophosphocholine | 0.000 | -0.006 | 0.005 | 8.88E-01 | -0.002 | -0.008 | 0.004 | 4.81E-01 | 0.001 | -0.003 | 0.006 | 4.93E-01 | 0.000 | -0.004 | 0.005 | 8.88E-01 |
| citicoline | 0.000 | -0.005 | 0.006 | 8.89E-01 | 0.001 | -0.005 | 0.006 | 7.94E-01 | 0.003 | -0.002 | 0.007 | 2.30E-01 | 0.005 | 0.000 | 0.009 | 5.78E-02 |
| uridinemonophosphate_ump | 0.000 | -0.005 | 0.006 | 9.08E-01 | 0.000 | -0.006 | 0.005 | 9.03E-01 | -0.001 | -0.006 | 0.003 | 5.31E-01 | -0.001 | -0.006 | 0.004 | 6.70E-01 |
| aspartate | 0.000 | -0.005 | 0.006 | 9.24E-01 | -0.001 | -0.006 | 0.005 | 8.02E-01 | -0.002 | -0.006 | 0.002 | 4.06E-01 | -0.005 | -0.009 | 0.000 | 4.01E-02 |
| phosphocholine | 0.000 | -0.006 | 0.005 | 9.26E-01 | -0.002 | -0.007 | 0.004 | 5.60E-01 | 0.000 | -0.004 | 0.004 | 9.84E-01 | -0.001 | -0.006 | 0.004 | 7.22E-01 |
| amino5_levulinic | 0.000 | -0.006 | 0.005 | 9.30E-01 | 0.000 | -0.005 | 0.006 | 8.80E-01 | -0.005 | -0.009 | -0.001 | 1.29E-02 | -0.002 | -0.006 | 0.003 | 4.67E-01 |
| acetyl_alanine | 0.000 | -0.005 | 0.006 | 9.39E-01 | 0.000 | -0.005 | 0.006 | 9.29E-01 | -0.005 | -0.008 | -0.001 | 1.93E-02 | -0.002 | -0.006 | 0.003 | 4.75E-01 |
| malic | 0.000 | -0.005 | 0.006 | 9.41E-01 | 0.001 | -0.005 | 0.006 | 8.23E-01 | -0.001 | -0.006 | 0.003 | 4.91E-01 | -0.003 | -0.007 | 0.002 | 2.78E-01 |
| aconitic | 0.000 | -0.005 | 0.006 | 9.46E-01 | 0.001 | -0.004 | 0.006 | 6.95E-01 | 0.002 | -0.002 | 0.006 | 3.43E-01 | -0.002 | -0.006 | 0.003 | 4.95E-01 |
| lysine | 0.000 | -0.006 | 0.005 | 9.56E-01 | 0.000 | -0.006 | 0.005 | 9.08E-01 | -0.003 | -0.008 | 0.001 | 1.07E-01 | -0.003 | -0.007 | 0.002 | 2.56E-01 |
| creatinine | 0.000 | -0.006 | 0.005 | 9.57E-01 | 0.000 | -0.006 | 0.005 | 9.89E-01 | 0.002 | -0.002 | 0.006 | 4.13E-01 | 0.001 | -0.003 | 0.006 | 5.71E-01 |
| ornithine | 0.000 | -0.005 | 0.005 | 9.64E-01 | -0.001 | -0.006 | 0.004 | 7.16E-01 | 0.007 | 0.003 | 0.011 | 1.60E-03 | 0.001 | -0.004 | 0.005 | 8.01E-01 |
| creatine | 0.000 | -0.005 | 0.005 | 9.67E-01 | 0.000 | -0.005 | 0.005 | 9.67E-01 | -0.005 | -0.009 | -0.001 | 6.99E-03 | -0.002 | -0.007 | 0.002 | 2.95E-01 |
| atorvastatin | 0.000 | -0.003 | 0.003 | 9.87E-01 | 0.000 | -0.003 | 0.003 | 9.13E-01 | 0.003 | 0.000 | 0.006 | 3.04E-02 | 0.002 | -0.001 | 0.005 | 2.81E-01 |
| c4butyrylcarnitine | 0.000 | -0.006 | 0.005 | 9.88E-01 | 0.000 | -0.006 | 0.005 | 9.86E-01 | 0.001 | -0.004 | 0.005 | 7.82E-01 | 0.001 | -0.003 | 0.006 | 5.64E-01 |
| citrulline | 0.000 | -0.005 | 0.005 | 9.90E-01 | -0.002 | -0.007 | 0.003 | 5.19E-01 | -0.002 | -0.006 | 0.003 | 4.77E-01 | -0.002 | -0.007 | 0.002 | 3.00E-01 |
